# Understanding the comorbidity between posttraumatic stress severity and coronary artery disease using genome-wide information and electronic health records

**DOI:** 10.1101/2022.03.04.22271901

**Authors:** Renato Polimanti, Frank R. Wendt, Gita A. Pathak, Daniel S. Tylee, Catherine Tcheandjieu, Austin T. Hilliard, Daniel F. Levey, Keyrun Adhikari, J. Michael Gaziano, Christopher J. O’Donnell, Themistocles L. Assimes, Murray B. Stein, Joel Gelernter

**Affiliations:** Department of Psychiatry, Yale School of Medicine, West Haven, CT, USA; VA CT Healthcare Center, West Haven, CT, USA; Department of Medicine, Stanford University School of Medicine, Stanford, CA, USA; VA Palo Alto Healthcare System, Palo Alto, CA, USA; Department of Medicine, VA Boston Healthcare System, Boston, MA, USA; Department of Medicine, Brigham and Women’s Hospital, Boston, MA, USA; Department of Medicine, Harvard Medical School, Boston, MA, USA; Department of Psychiatry, University of California San Diego, La Jolla, CA, USA; VA San Diego Healthcare System, Psychiatry Service, San Diego, CA, USA; Herbert Wertheim School of Public Health and Human Longevity Science, University of California San Diego, La Jolla, CA, USA; Departments of Genetics and of Neuroscience, Yale School of Medicine, New Haven, CT, USA

**Keywords:** genetics, causal inference, comorbidity, cardiovascular disease, trauma

## Abstract

**Background:** The association between coronary artery disease (CAD) and posttraumatic stress disorder (PTSD) contributes to the high morbidity and mortality observed among affected individuals. To understand the dynamics underlying PTSD-CAD comorbidity, we conducted a genetically-informed causal inference analysis using large-scale genome-wide association (GWA) statistics and follow-up analysis using electronic health records (EHR) and PTSD Checklist (PCL-17 or PCL-6) assessments available from the Million Veteran Program (MVP) and the UK Biobank (UKB), respectively.

**Methods:** We used GWA statistics from MVP, UKB, the Psychiatric Genomics Consortium, and the CARDIoGRAMplusC4D Consortium to perform a bidirectional, two-sample Mendelian randomization (MR) analysis to assess cause-effect relationships between CAD and PTSD. We also conducted a pleiotropic meta-analysis to investigate loci with concordant vs. discordant effects between the traits investigated. Leveraging individual-level information derived from MVP and UKB EHRs, we assessed longitudinal changes in the association between CAD and posttraumatic stress severity.

**Findings:** We observed a genetic correlation of CAD with PTSD case-control and quantitative outcomes, ranging from 0.18 to 0.32. Our two-sample MR showed a significant bidirectional relationship between CAD and PTSD symptom severity. Genetically-determined PCL-17 total score was associated with increased CAD risk (odds ratio=1.04; 95% confidence interval, 95%CI=1.01-1.06). Conversely, CAD genetic liability was associated with reduced PCL-17 total score (beta=-0.42; 95%CI=-0.04 – -0.81). These estimates were consistent across datasets and were not affected by heterogeneity or horizontal pleiotropy. The pleiotropic meta-analysis between PCL-17 and CAD identified loci with concordant effect enriched for platelet amyloid precursor protein pathway (p=2.97×10^−7^) and negative regulation of astrocyte activation (p=2.48×10^−6^) while discordant-effect loci were enriched for biological processed related lipid metabolism (e.g., triglyceride-rich lipoprotein particle clearance, p=1.61×10^−10^). The EHR-based follow-up analysis highlighted that earlier CAD diagnosis is associated with increased PCL-total score later in life, while lower PCL total score was associated with increased risk of a later CAD diagnosis (Mann-Kendall trend test: MVP tau=0.932, p<2×10^−16^; UKB tau=0.376, p=0.005)

**Interpretation:** Our results highlight a complicated relationship between PTSD and CAD that may be affected by the long-term consequences of CAD on the mental health of the individuals affected.

**Funding:** This research was supported by funding from the VA Cooperative Studies Program (CSP, no. CSP575B) and the Veterans Affairs Office of Research and Development MVP (grant nos. MVP000 and VA Merit MVP025).

## INTRODUCTION

Posttraumatic stress disorder (PTSD) is a psychiatric illness associated with long-term impairment following severe traumatic experiences^1^. PTSD lifetime prevalence can be as high as 30% in high-risk groups (e.g., individuals from war-exposed regions)^2^. In addition to disorders of mental health, somatic comorbidities greatly contribute to the increased mortality observed in PTSD cases^3^, and in particular, the incidence of cardiovascular diseases (CVD) is a major contributor to PTSD premature mortality. Several studies have applied different approaches to investigate the underlying dynamics linking these illnesses^4^, but there is no clear understanding of whether PTSD-CVD associations are due to causal effects or to shared pathogenic processes.

A major challenge to disentangle this complex relationship is to model the longitudinal changes that lead to PTSD-CVD comorbidity. It is of great importance to understand these relationships because they are fundamental to many of the questions we wish to answer, including: does CVD cause PTSD? And, does having PTSD increase future risk of CVD? In recent years, resources generated by the analysis of massive cohorts have revolutionized our ability to investigate the dynamics linking mental and physical health. In particular, genome-wide datasets are permitting investigators to assess evidence of possible cause-effect relationships. For instance, genetic associations can be used as instrumental variables in Mendelian randomization (MR) analyses to test the causal effect of exposures on outcomes of interest^5^. Genetically-informed causal inference analyses can be used to investigate cause-effect hypotheses that may otherwise not be feasible to test in randomized trials due to practical or ethical considerations. For PTSD, MR has been used to investigate causal hypotheses with respect to psychiatric comorbidities^6,7^, obesity-related traits^8,9^, respiratory outcomes^10^, tobacco smoking^11^, inflammatory biomarkers^12^, sleeping patterns^13^, and socioeconomic factors^14-16^. These MR studies support a complex network of cause-effect relationships involving PTSD. With respect to CVD outcomes, no genetically-informed causal inference analysis has tested a hypothesis to explain the comorbidity with PTSD. However, a recent study showed that PTSD symptoms are genetically correlated with hematologic and cardiometabolic traits^17^.

To provide novel insights into the dynamics underlying PTSD-CVD comorbidity, we leveraged genome-wide information from the Million Veteran Program (MVP)^18^, the Psychiatric Genomics Consortium (PGC)^19^, the CARDIoGRAMplusC4D Consortium^20^, and the UK Biobank (UKB)^21^. Our primary analysis was focused on coronary artery disease (CAD), for which there is strong evidence for association with PTSD^22,23^. For PTSD, we analyzed genetic instruments derived from a binary case-control definition and from quantitative assessment of PTSD symptom clusters. To follow up our genetically-informed causal inference analysis, we investigated the electronic health records (EHR) available for 319,036 MVP participants and 155,817 UKB participants who were also assessed with PTSD Checklist instruments (PCL; for MVP the 17-item checklist, PCL-17; for UKB the six-item checklist, PCL-6)^24^.

## METHODS

### Study Design

This study was conducted using genome-wide association (GWA) statistics generated using data from MVP, UKB, PGC, and the CARDIoGRAMplusC4D Consortium and individual-level information from MVP and UKB electronic health records (EHR) and PCL assessments. The use of GWA statistics (i.e., previously collected, deidentified, aggregated data) did not require institutional review board approval. The use of UKB individual-level data has been conducted through the application reference no. 58146. UKB has approval from the North West Multi-center Research Ethics Committee (MREC) as a Research Tissue Bank (RTB) approval. This approval means that researchers do not require separate ethical clearance and can operate under the RTB approval. The use of MVP individual-level data was conducted under project CSP575b. MVP is approved by the VA Central IRB and project CSP575b was also approved by local VA IRBs in Boston, San Diego, and West Haven.

We implemented a multi-step analytic design to identify possible causal effects underlying PTSD-CAD comorbidity. Initially, we used GWA statistics to investigate the genetic correlations among CAD, PTSD, and related outcomes. Then, we conducted a polygenic risk score (PRS) analysis to define genetic instruments for the two-sample MR analysis. We followed the STROBE (STrengthening the Reporting of Observational studies in Epidemiology) guidelines^25^ to design and report our MR analyses and the related sensitivity tests. To verify the MR findings with an alternative genetically-informed causal inference method, we used the latent causal variable (LCV) approach^26^. To explore further the dynamics linking CAD and PTSD, we investigated CAD-related EHR diagnoses of MVP and UKB participants who were also assessed with PCL instruments. Specifically, we tested the association between CAD and PTSD severity stratifying our sample by CAD onset with respect to the timing of the PCL assessment (i.e., CAD onset before PCL assessment, CAD onset the same year as PCL assessment, CAD onset after PCL assessment). Additionally, within CAD cases, we also tested the relationship of PTSD severity with differential time to onset of CAD and PCL assessment

### Cohorts

This study investigated CAD-PTSD comorbidity using genetic and phenotypic information derived from multiple cohorts. This permitted us to verify whether the associations detected were due to cohort-specific characteristics or whether they could be generalized across different study populations. Specifically, we analyzed datasets generated from biobanks (i.e., MVP and UKB) and from GWAS (GWA study) meta-analyses of cohorts assessed with different diagnostic instruments (i.e., PGC and CARDIoGRAMplusC4D Consortium).

#### Million Veteran Program

This is a biobank representative of the US Veterans that are active users of the Veterans Health Administration (VHA) healthcare system^27^. To date, more than 825,000 individuals were enrolled in MVP and high-quality genotype information is available for >650,000 MVP participants^18^. In our study, we investigated MVP genome-wide information and EHRs related to PTSD and CAD. We used GWA statistics from a recent PTSD GWAS conducted in MVP^28^ that analyzed an EHR-validated algorithmic PTSD diagnosis ^29^ and PCL-17-derived quantitative phenotypes related to PTSD symptom clusters (re-experiencing, REX; avoidance, AVOID; hyperarousal, HYPER; and overall severity score, PCL-17)^28^. We included GWA statistics generated from 36,301 cases and 178,107 controls of European descent for the PTSD diagnosis and from 186,689 individuals of European descent for the PCL-17-derived quantitative phenotypes. For CAD, we used GWA statistics from a CAD GWAS conducted in 293,470 MVP participants of European descent. In our phenotype-only follow-up analysis, we investigated PTSD information derived from the PCL-17 assessment and CAD information derived from VHA EHRs. As part of the MVP lifestyle survey, MVP participants completed a PCL-17 assessment, which asks respondents to report the extent to which they had been affected in the previous month by symptoms in response to stressful life experiences. To derive CAD diagnosis from VHA EHR, we used the ICD (International Classification of Diseases) codes used by the CARDIoGRAMplusC4D Consortium^20^. Briefly, CAD was defined as myocardial infarction, chronic ischemic heart disease, or angina (ICD9: 410-413, 414.0, 414.8, 414.9; ICD10: I20, I21-I24, I25.1, I25.2, I25.5-I25.9). Exclusion criteria were aneurysm and atherosclerotic cardiovascular disease (ICD9 414.1 and ICD 10 I25.0, I25.3, I25.4). Through this CAD definition, we identified 73,074 cases and 245,962 controls that completed the PCL-17 assessment.

#### UK Biobank

This is an open-access resource available to investigate a wide range of traits including serious and life-threatening illnesses^21^. More than 500,000 UKB participants were assessed through detailed web-based questionnaires on their diet, cognitive function, work history, health status, and other relevant phenotypes. Additionally, data are derived from EHRs. UKB contributed to the PGC and CARDIoGRAMplusC4D GWAS meta-analyses investigated in our study. A description of UKB contribution to these datasets is provided in the subsequent paragraphs. As part of the online-mental health questionnaire^30^, 157,366 participants were also assessed for items included in the PCL-6 assessment. Similar to the CARDIoGRAMplusC4D Consortium^20^, we defined CAD cases leveraging UKB information derived from questionnaires and EHRs. Based on the questionnaires, we defined CAD cases as UKB participants having ‘vascular/heart problems diagnosed by doctor’, ‘non-cancer illnesses that self-reported as angina or heart attack’, or self-reported surgery including ‘percutaneous transluminal coronary angioplasty’, ‘coronary artery bypass grafting’, or ‘triple heart bypass’. Through UKB EHRs, we defined CAD cases as participants reporting codes related to infarction, percutaneous transluminal coronary angioplasty, coronary artery bypass grafting, chronic ischemic heart disease, or angina (ICD9: 410-413, 414.0, 414.8, 414.9; ICD10: I20, I21-I24, I25.1, I25.2, I25.5-I25.9; OPCS-4: K40–K46; K49, K50.1, K75). Exclusion criteria were aneurysm and atherosclerotic cardiovascular disease (ICD9 414.1 and ICD 10 I25.0, I25.3, I25.4). Through this CAD definition, we identified 8,942 cases and 146,875 controls that completed the PCL-6 assessment.

#### Psychiatric Genomics Consortium

This is an international collaborative effort focused on gene discovery of psychiatric traits and disorder^19^.We used GWA statistics generated by the PGC-PTSD workgroup, which conducted GWAS meta-analysis across 60 different PTSD studies, ranging from clinically deeply characterized, small patient groups to large cohorts with self-reported PTSD symptoms^31^. We analyzed two datasets: PGC-PTSD version 1.5 including 12,823 cases 35,648 controls of European descent; PGC-PTSD version 2 (i.e., PGC-PTSD version 1.5 plus UKB) including 23,212 cases 151,447 controls of European descent.

#### CARDIoGRAMplusC4D Consortium

This effort combines data from multiple large-scale genetic studies to identify risk loci for coronary artery disease and myocardial infarction. We analyzed two CARDIoGRAMplusC4D datasets: a GWAS meta-analysis of 48 studies including 60,801 cases and 123,504 controls^32^ and a follow-up study added UKB to a novel meta-analysis reaching a total of 79,602 cases and 261,418 controls^20^.

#### Other Datasets

To explore further the comorbidity of PTSD across CVDs, we investigated additional traits, including blood pressure, heart failure, PR interval, and resting heart rate. For blood pressure, we analyzed GWA statistics related to systolic blood pressure (N=738,170)^33^, diastolic blood pressure (N=757,601)^33^, pulse pressure (N=738,170)^33^, and self-reported high blood pressure (144,793 case and 313,761 controls)^34^. For heart failure, the GWA statistics used in the present study were generated from 47,309 cases and 930,014 controls^35^. For resting heart rate, we used GWA statistics derived from 458,969 individuals^34^.

### Data Analysis

#### Effect size distribution of genetic effects in PTSD and CAD phenotypes

To investigate the genetic architecture of PTSD and CAD traits, we applied GENESIS (GENetic Effect-Size distribution Inference from Summary-level data) package^36^.

#### Genetic Correlation Analysis

Initially, we assessed the informativeness of the GWAS datasets analyzed using the linkage disequilibrium (LD) score regression method^37^. This permitted us to estimate the genetic correlation (rg) among them. Considering genetic correlations observed, we evaluated the consistency within PTSD-related traits assessed in different cohorts (MVP and PGC) and within CAD-related traits assessed in different cohorts (MVP and CARDIoGRAMplusC4D Consortium). We then studied the genetic correlation among PTSD-related traits, CAD, and traits related to traumatic experiences and social support.

#### Polygenic Risk Scoring

Considering those cross-phenotype genetic correlations that survived Bonferroni multiple testing correction, we conducted a PRS analysis based only on GWA statistics. We used the gtx R package incorporated in PRSice software^38^ to calculate an approximate estimate of the explained variance from a multivariate regression model^39^. We defined PRS considering a genome-wide significance threshold (P<5×10^−8^) and applying a clumping procedure with an LD cutoff of R^2^ = 0.001 within a 10,000-kilobase window, excluding the major histocompatibility complex region of the genome because of its complex LD structure. The European samples from the 1000 Genomes Project were used as the LD reference panel^40^.

#### Genetically-informed Causal Inference Analysis

We used multiple GWA statistics to identify putative causal effects underlying PTSD-CAD comorbidity. Similar to previous studies^9,12,16^, we applied a multi-step analytic design, assessing the informativeness of the datasets, defining the genetic instruments via PRS analysis, and conducting causal inference analyses independently via MR and LCV approaches.

Considering genetic instruments that survived multiple testing correction in the PRS analysis, we conducted a two-sample MR analysis using the TwoSampleMR R package^41^. Since different MR methods have different sensitivities to different potential confounders, accommodate different scenarios, and vary in their statistical efficiency^41^, we considered a range of MR methods. The primary analysis was conducted considering a random-effects inverse-variance weighted (IVW) method^41^. The secondary MR methods included MR Egger^42^, simple mode^43^, weighted median^44^, and weighted mode^43^. We conducted multiple MR sensitivity analyses to exclude possible biases (horizontal pleiotropy, i.e. the variants included in the genetic instrument have an effect on disease outside their effects on the exposure in MR^45^) under different scenarios in the MR estimates. These included the IVW-heterogeneity test^46^, the MR-Egger intercept^42^, the MR-Robust Adjusted Profile Score (MR-RAPS) overdispersion test^47^, and the MR–Pleiotropy Residual Sum and Outlier (MR-PRESSO) global test^48^. Finally, a leave-one-out analysis was conducted to identify potential outliers among the variants included in the genetic instruments tested. To validate the MR findings with an alternative method for genetically-informed causal inference, we used the LCV approach^26^. This postulates that the genetic correlation between two traits can be mediated by a latent variable that has a causal effect on the traits tested^26^. LCV estimates the genetic causality proportion (GCP) using z-score converted per-variant effects and regression weights from GWA statistics^26^.

#### Pleiotropic Meta-Analysis

We leveraged ASSET (Association Analysis based on Subsets) approach^49^ to identify loci with concordant effect directions phenotypes and loci with discordant effect directions between PCL-17 (MVP) and CAD (UKB-CARDIoGRAMplusC4D). The ASSET results were annotated using Multi-marker Analysis of GenoMic Annotation (MAGMA) ^50^ implemented in FUMA^51^ and tested for enrichment with respect to gene sets available from the Molecular Signatures Database (MSigDB)^52^.

#### EHR-based Follow-Up Analysis

We used individual-level EHR and PCL data available from MVP and UKB cohorts to explore further the association between CAD and PTSD symptom severity. Specifically, we assessed the association of CAD with PCL total score, stratifying the analysis by the difference between the time of the PCL assessment with respect to the time of the first CAD diagnosis. As described in the section *Cohorts*, MVP participants were screened using the PCL-17 assessment and CAD diagnosis was based on the ICD codes defined by the CARDIoGRAMplusC4D Consortium. UKB participants were screened using the PCL-6 assessment and CAD diagnosis was based on the ICD and OPCS-4 codes and on self-reported information using the same approach previously applied for this cohort by the CARDIoGRAMplusC4D Consortium^20^. A linear regression analysis was conducted to test the association of CAD with PCL total score. In MVP analysis, we included age, sex, income, self-reported ethnicity, and self-reported racial background as covariates. In UKB analysis, we included age, sex, Townsend deprivation index (a measure of material deprivation^53^), and self-reported ethnic background. Within CAD cases, we also tested the association of PCL total score with the difference between the time of the PCL assessment and the time of the first CAD diagnosis. This analysis was conducted considering a linear regression model using the same sets of covariates described above. We also used the Mann-Kendall trend test to verify whether there is a statistically significant trend in the changes of PCL score across CAD cases depending on the difference in time between CAD diagnosis and PCL assessment.

## RESULTS

### Genetic Correlation Analysis

To investigate the dynamics linking PTSD and CAD, we used multiple genome-wide datasets generated from different cohorts. Among them, we observed a high genetic correlation for both PTSD/PCL and CAD (rg>0.95; Supplemental Table 1). Comparing their genetic architectures, PTSD traits had a higher degree of polygenicity (i.e., higher number of variants with small individual effect) than CAD (Supplemental Figure 1). However, there were differences between binary and quantitative traits (i.e., PTSD binary phenotype is more polygenic than PTSD quantitative phenotypes) and cohort-specific differences (e.g., CAD appears less polygenic in the CARDIoGRAMplusC4D GWAS meta-analysis than in the GWAS meta-analysis including both UKB and CARDIoGRAMplusC4D; Supplemental Figure 1).

For three CAD datasets investigated (i.e., CARDIoGRAMplusC4D, UKB-CARDIoGRAMplusC4D, and MVP), the genetic correlation between CAD and PTSD traits ranged from 0.18 to 0.32 (Figure 1A; Supplemental Table 2). We did not observe dataset-specific differences, but the more extreme results were mainly due to a higher degree of uncertainty in the less powered PTSD datasets. The more informative PTSD datasets converged around the most significant estimate (i.e., rg=0.22, p=1.36×10^−13^; Supplemental Table 2).

**Figure 1:**
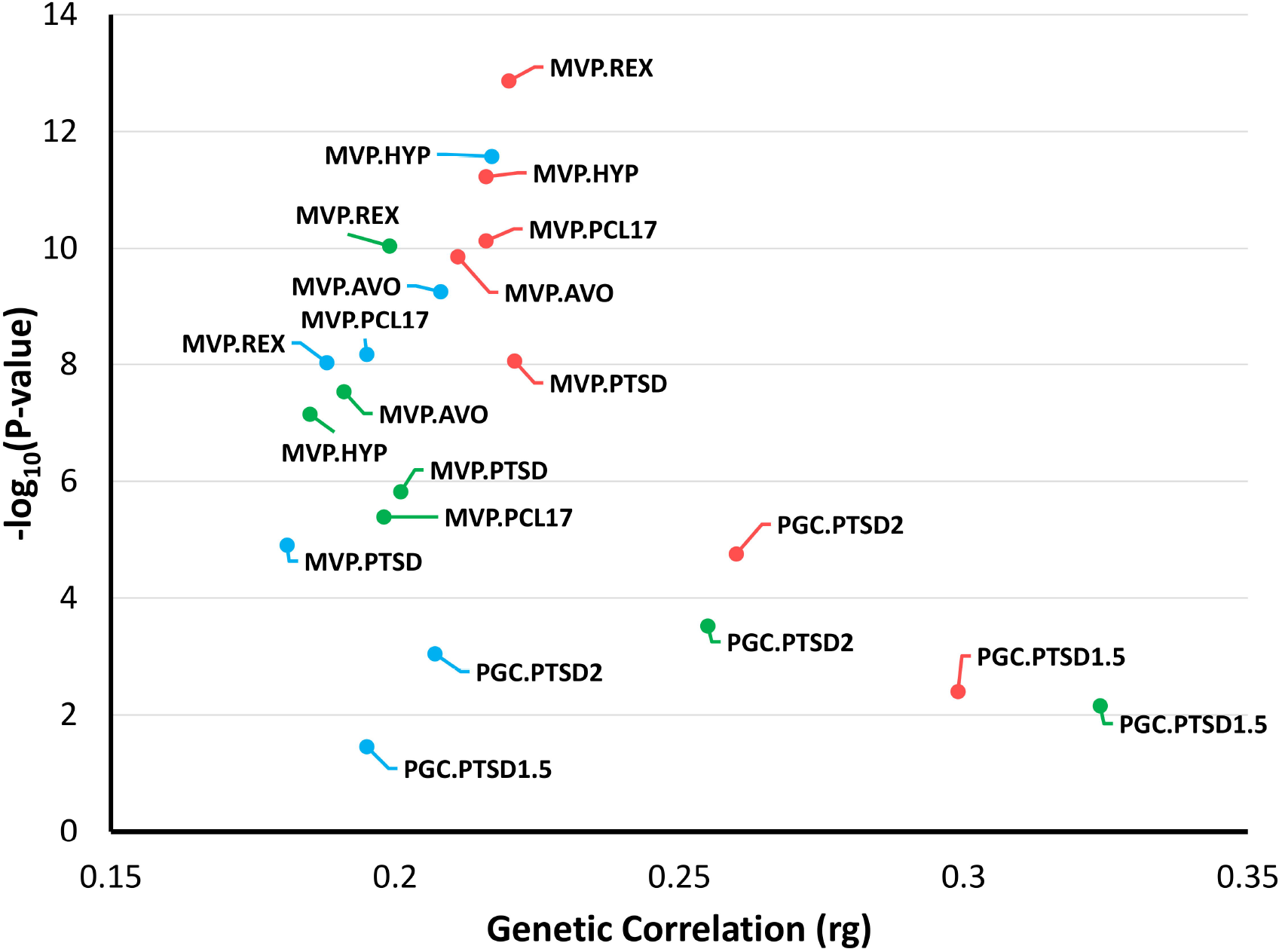
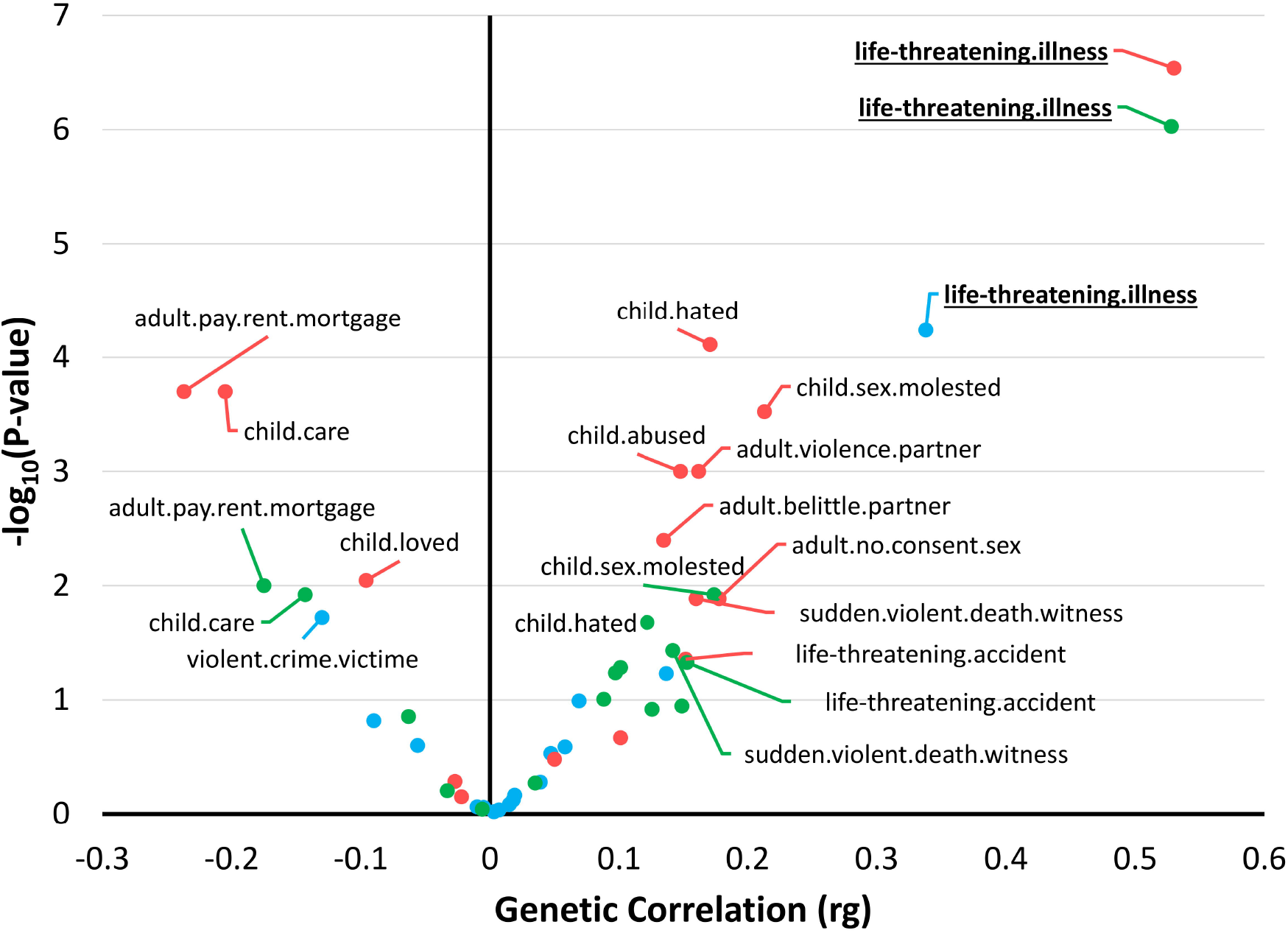
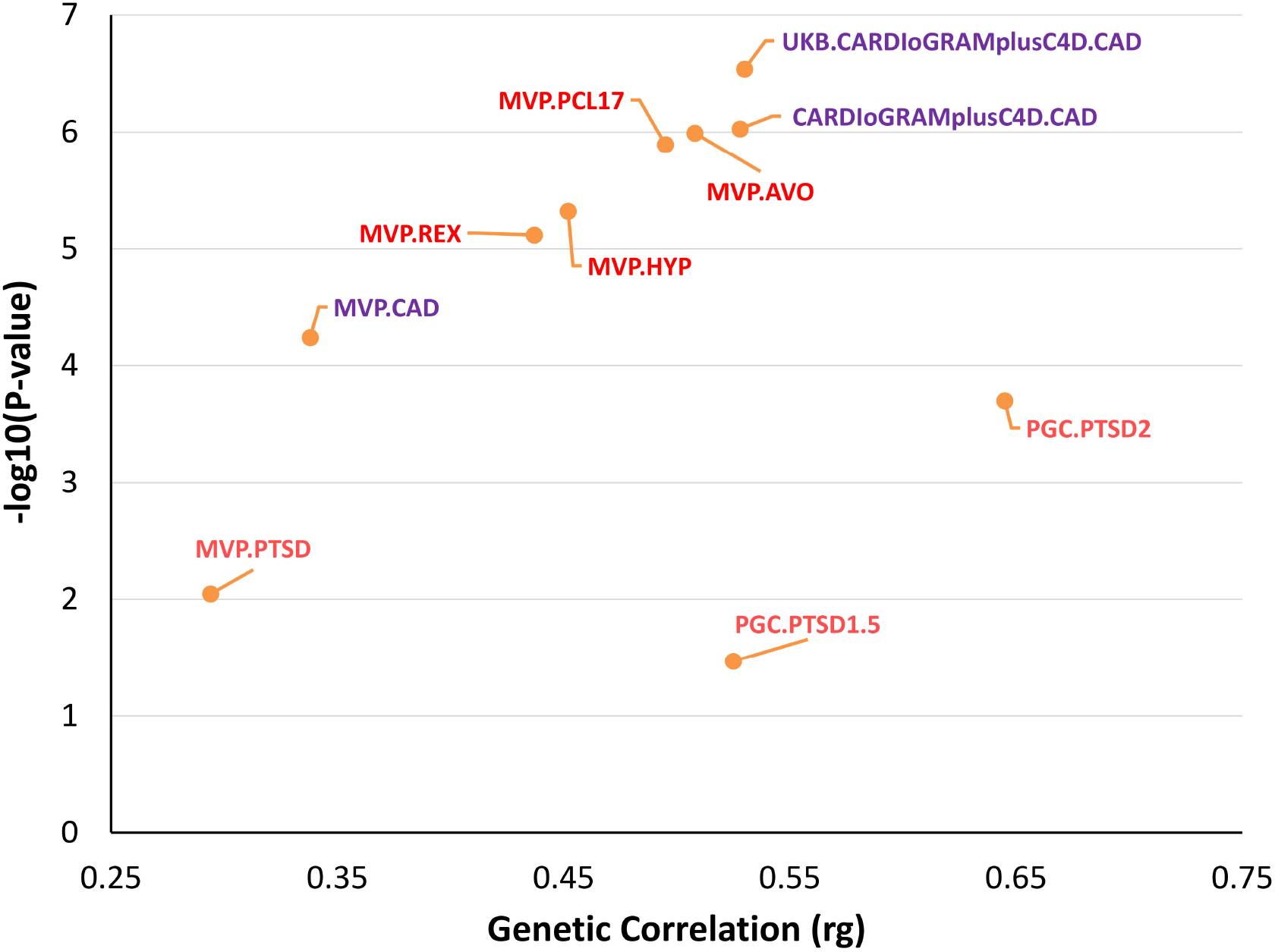
**A**) Genetic correlation (rg) between CAD and PTSD traits. Details are reported in Supplemental Table 2. **B**) Genetic correlation of CAD with traits related to traumatic events and social support. Details are reported in Supplemental Table 3. Labels are reported for genetic correlation surviving multiple testing correction. Underlined bolded labels are reported for genetic correlation surviving multiple testing correction across the three CAD datasets tested. **C**) Genetic correlation of CAD and PTSD with “Been diagnosed with a life-threatening illness”. Details are reported in Supplemental Table 4. In panels **A** and **B**, CAD datasets are color-coded as light blue for MVP, red for UKB-CardioGRAMplusC4D meta-analysis, and green for CardioGRAMplusC4D meta-analysis.

To explore further the relationship between PTSD and CAD, we investigated the genetic correlation of CAD with 16 UKB-derived traits related to traumatic events and social support (Figure 1B; Supplemental Table 3). Considering a Bonferroni correction accounting for the number of traits tested, only “Been diagnosed with a life-threatening illness” (UKB Data-Field: 20528) showed statistically significant genetic correlation with the CAD datasets investigated (CARDIoGRAMplusC4D: rg=0.53, p=9.37×10^−7^; UKB-CARDIoGRAMplusC4D: rg=0.53, p=2.89×10^−7^; MVP: rg=0.34, p=5.78×10^−5^). Although MVP CAD showed a lower genetic correlation with “Been diagnosed with a life-threatening illness” than the other two CAD datasets investigated, this difference was not statistically significant (p>0.05). Conversely, we observed strong dataset-specific differences for other trauma-related traits investigated. Considering the UKB-CARDIoGRAMplusC4D dataset, we identified significant genetic correlations of CAD with six other phenotypes, but none of these survived multiple testing correction in the other two CAD datasets. For example, the strongest UKB-CARDIoGRAMplusC4D genetic correlation was observed for “Felt hated by family member as a child” (UKB-CARDIoGRAMplusC4D: rg=0.17, p=7.72×10^−5^), but the other CAD datasets did not show the same statistical power or magnitude of genetic correlation (CARDIoGRAMplusC4D: rg=0.12, p=0.021; MVP: rg=0.069, p=0.102). Although rg estimates calculated via LD score regression are not biased by sample overlap^37^, the characteristics of the UKB cohort due to recruitment and assessment strategies^54^ may have inflated the genetic correlations between CAD assessed in the UKB-CARDIoGRAMplusC4D meta-analysis and traits related to traumatic events and social support assessed in the UKB. For the variable “Been diagnosed with a life-threatening illness”, CAD and PTSD have a comparable genetic correlation across the datasets investigated (Figure 1C; Supplemental Table 4).

### Genetically-informed Causal Inference Analysis

To understand possible cause-effect relationships underlying the PTSD-CAD genetic correlation, we conducted a bidirectional MR analysis. Similar to previous studies^16^, we defined informative genetic instruments conducting a cross-phenotype PRS analysis based on variants reaching genome-wide significance (p<5×10^−8^) in the exposure GWAS. Because the two-sample MR approach can be biased by sample overlap^55^, we selected non-overlapping exposure and outcome datasets (see *Cohorts* description) and prioritized the largest sample size when multiple datasets were available. Applying a Bonferroni multiple testing correction (p=0.005), we identified seven significant PRS associations, observing bidirectional relationships between CAD and PTSD traits (Supplemental Table 5). As shown in the genetic correlation analysis (Figure 1A), datasets based on PTSD quantitative traits are statistically more powerful than the ones based on PTSD binary definition although they have an almost perfect genetic correlation with each other (rg>0.95; Supplemental Table 1). Accordingly, we investigated the cause-effect relationship between CAD (UKB-CARDIoGRAMplusC4D) and PCL-17 total score (MVP) as the primary test in our two-sample MR analysis. While genetically-determined PCL-17 total score was associated with increased CAD risk (IVW odds ratio=1.04, p=2.56×10^−3^; Figure 2A), genetically-determined CAD was associated with decreased PCL-17 total score (IVW beta=-0.42, p=0.029). These estimates were consistent across different MR methods and no evidence of horizontal pleiotropy was detected via the MR-Egger intercept (Supplemental Table 6). However, the CAD→PCL-17 association was characterized by strong heterogeneity (IVW heterogeneity test: Q=89.4, p=2.45×10^−7^; MR-RAPS global test: RSSobs=110.9, p<10^−4^). We conducted a leave-one-out (LOO) analysis to identify the variants contributing to the heterogeneity within the CAD instrumental variable (Supplemental Table 7). After removing the outliers identified in the LOO analysis, we confirmed that increased CAD genetic liability is associated with reduced PCL-17 score (IVW beta=-0.50, p=2.61×10^−4^; Figure 2B, Supplemental Table 8) in the absence of heterogeneity (IVW heterogeneity test: Q=36.8, p=0.121; MR-RAPS global test: RSSobs=39.6, p=0.137). We observed similar effects of CAD across PTSD symptom cluster severity (i.e., AVOID, HYPER, and REX; Supplemental Table 9). These estimates were characterized by strong heterogeneity, but the effects were confirmed after the removal of outlier variants from the instrumental variables (Supplemental Table 10).

**Figure 2:**
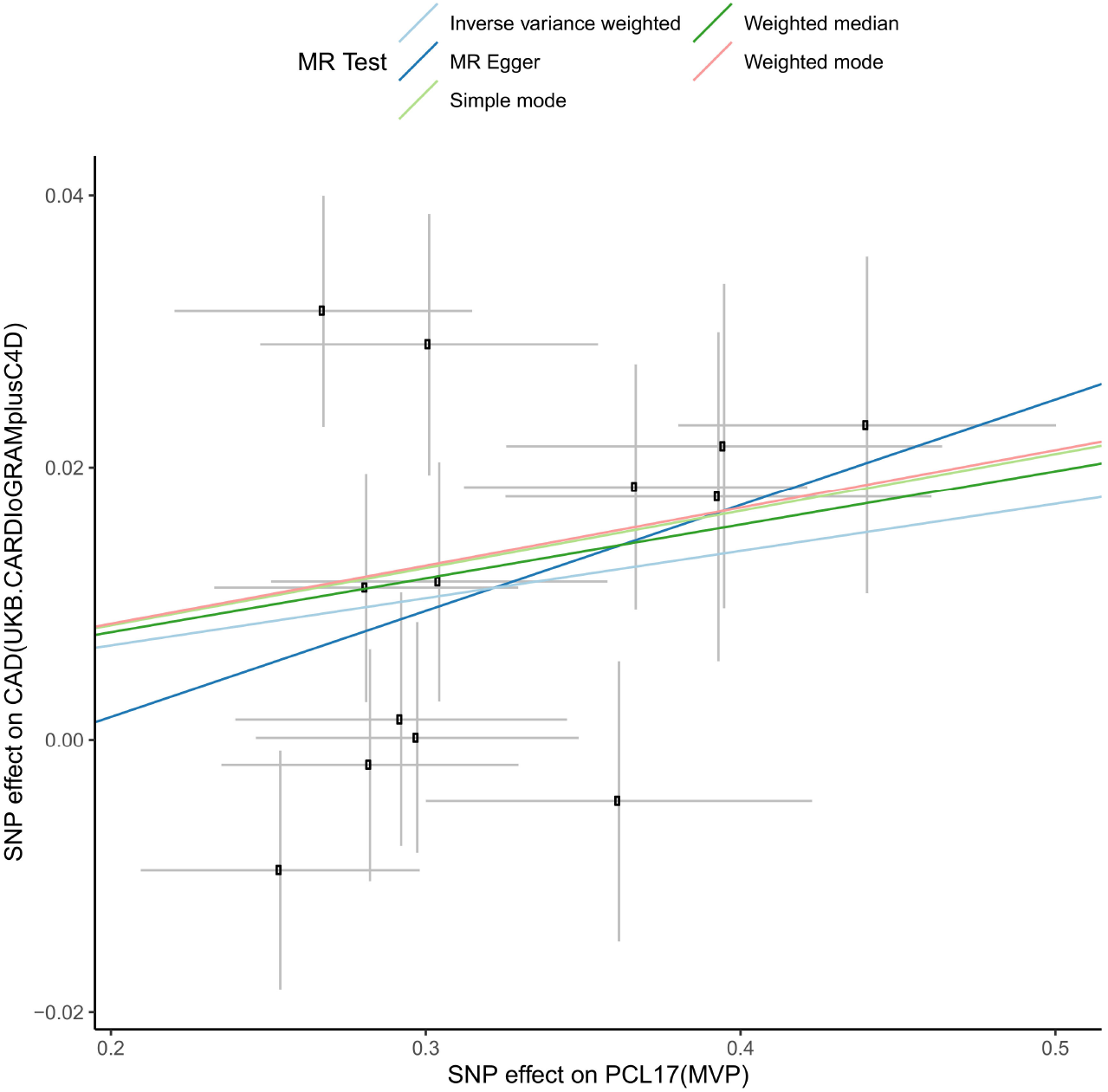
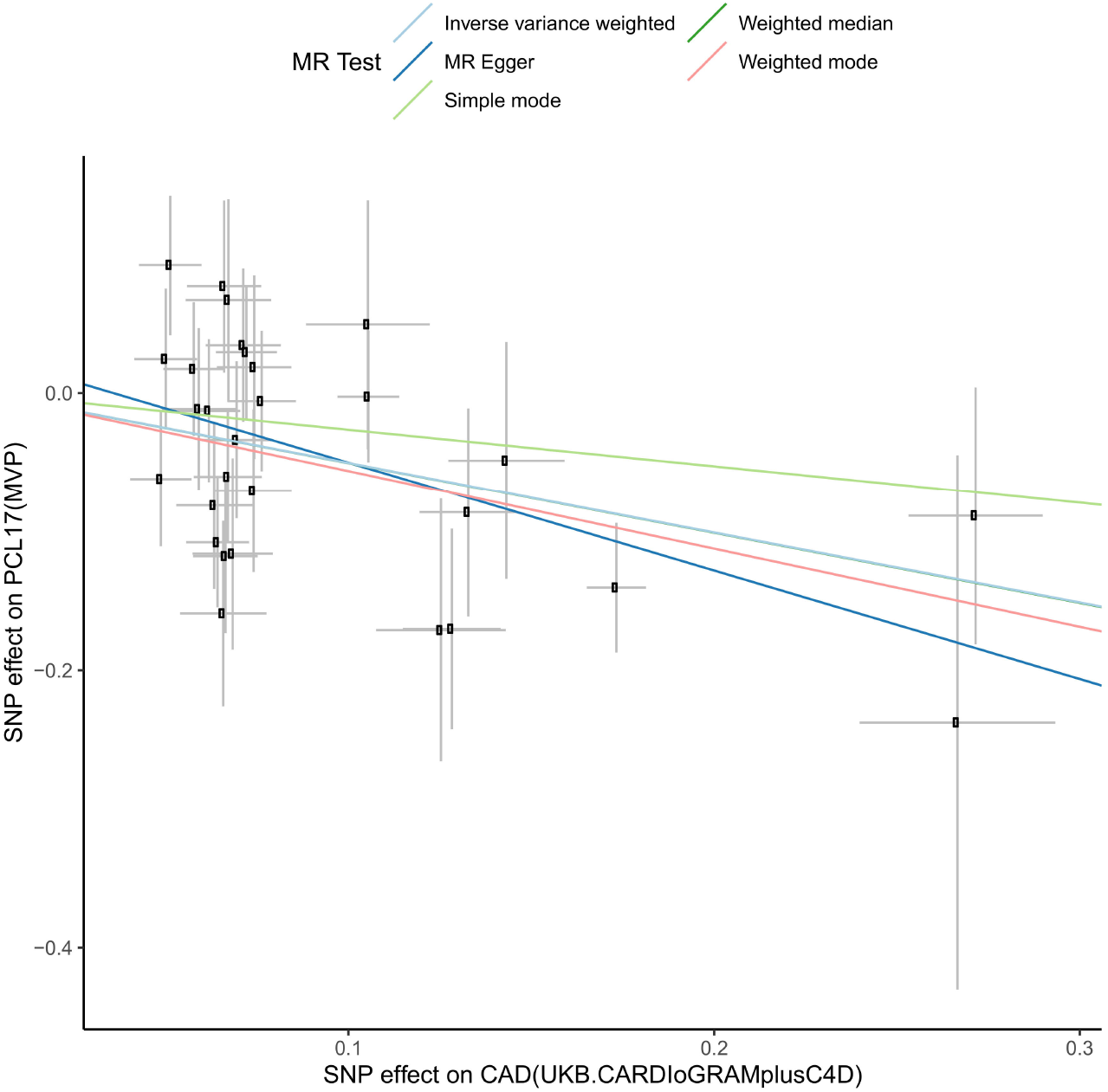
**A**) SNP-exposure (CAD associations, logOR) and SNP-outcome (PCL17 associations, beta) coefficients. **B**) SNP-exposure (PCL17 associations, beta) and SNP-outcome (CAD associations, logOR) coefficients. Error bars (95% CIs) are reported for each association. Slopes are reported for multiple MR tests.

To evaluate whether the CAD→PCL-17 effect was due to characteristics of the datasets investigated (UKB-CARDIoGRAMplusC4D CAD GWAS and MVP PCL-17 GWAS), we re-tested the association using the other datasets available. Using the CARDIoGRAMplusC4D GWAS (not including UKB) as the exposure dataset, we observed the same pattern of associations across the PCL-17-derived traits (total score: beta=-0.44, p=2.48×10^−3^; AVOID: beta=-0.15, p=8.71×10^−3^; HYPER: beta=-0.14, p=1.91×10^−3^; REX: beta=-0.13, p=1.24×10^−3^; Supplemental Table 11). Similar to the analyses conducted using the UKB-CARDIoGRAMplusC4D GWAS, there was evidence of significant heterogeneity in these associations (Supplemental Table 11), but the estimates were confirmed after the removal of the variant outliers (Supplemental Table 12). With regard to PCL-17, there is no large-scale GWAS conducted in a cohort other than MVP. However, there are two GWAS datasets informative for PTSD severity: a PCL-6 GWAS conducted in the UKB cohort and a PGC GWAS of a PTSD quantitative phenotype combining UKB PCL-6 with PTSD severity information from other cohorts^56^. To avoid sample overlap bias, the MR analyses were conducted testing CARDIoGRAMplusC4D CAD → UKB PCL-6 and ii) MVP CAD → PGC PTSD quantitative phenotype. The effects observed from these analyses were null with inconsistent effect direction across MR methods in both cases (Supplemental Table 13). Unlike the high genetic correlation among PTSD traits assessed in different cohorts (rg>0.95; Supplemental Table 1), the genetic correlation of MVP PCL-17 was lower with respect to UKB PCL-6 (rg=0.81, p=6.26×10^−75^) and PGC PTSD quantitative phenotype (rg=0.84, p=2.5×10^−75^).

To evaluate whether the effect observed was specific to CAD, we tested other cardiovascular outcomes and risk factors (see Methods). We identified significant genetic correlation of MVP PCL-17 total score with heart failure (rg=0.24, p=9.45×10^−7^), self-reported high blood pressure (rg=0.23, p=5.86×10^−16^), and resting heart rate (rg=0.15, p=1.1×10^−10^). Among them, we observed significant PRS association with MVP PCL-17 only for heart failure (Supplemental Table 14). Our MR analysis showed that increased risk of genetically-determined heart failure is associated with lower PCL-17 (beta=-1.35, p=1.71×10^−4^). This effect was consistent across different MR methods and no evidence of heterogeneity and horizontal pleiotropy was observed (Supplemental Table 15).

To explore further the putative cause-effect relationships identified in our MR analysis, we applied the LCV approach. This is an alternative method to perform causal inference analyses modeling genetic effects^26^. LCV analysis did not identify a statistically significant genetic causality proportion between these traits (Supplemental Table 16).

### Pleiotropic Meta-Analysis

Testing for concordant and discordant genetic association between PCL-17 (MVP) and CAD (UKB-CARDIoGRAMplusC4D), we identified seven variants with concordant effects and ten variants with discordant effects that reached genome-wide significance (p<5×10^−8^) in the pleiotropic meta-analysis (Supplemental Table 17). To investigate the underlying mechanisms of concordant and discordant genetic effects, we conducted an enrichment analysis for molecular pathways and gene ontologies (GO). After applying a Bonferroni multiple testing correction (p=3.25×10^−6^), we identified two processes that were enriched for loci with concordant effects between PCL17 and CAD: “*Platelet Amyloid Precursor Protein (APP) Pathway*” (MSigDB: M6487; beta=1.53, p=2.97×10^−7^) and “*Negative Regulation of Astrocyte Activation*” (GO:0061889; beta=1.51, p=2.48×10^−6^). Seven enrichments were identified with loci with discordant effect: “*Triglyceride-Rich Lipoprotein Particle Clearance*” (GO:0071830; beta= 2.32, p=1.61×10^−10^), “*Chylomicron Clearance*” (MSigDB: M27845; beta=2.99, p=8.92×10^−9^); “*Low-Density Lipoprotein (LDL) Pathway during Atherogenesis*” (MSigDB: M22051; beta=2.45, p=3.18×10^−8^); “*Protein-Lipid Complex Assembly*” (GO:0065005; beta=0.94, p=7.48×10^−8^), “*Phosphatidylcholine Catabolic Process*” (GO:0034638; beta=1.58, p=2×10^−6^); “*Tumor Suppressor Arf Inhibits Ribosomal Biogenesis*” (MSigDB: M11358; beta=1.08, p=1.62×10^−6^); “*Plasma Lipoprotein Assembly*” (MSigDB: M27841; beta=1.31; p=3.06×10^−6^).

### EHR-based Follow-Up Analysis

To disentangle the possible dynamics underlying the effects detected using GWA statistics, we used individual-level EHR and PCL information from MVP and UKB. Specifically, we considered the temporal relationship between CAD (status and time of the diagnosis) and PCL (total score and time of the assessment). Stratifying CAD cases for whether the disease diagnosis was before, after, or the same year of the PCL assessment, we observed positive associations of CAD with PCL total score across all comparison groups in both MVP and UKB (Table 1). Within CAD cases, we also investigated whether the time difference between CAD onset and PCL assessment (Supplemental Figures 2 and 3) was associated with PCL total score. In both MVP and UKB, there was a significant trend where CAD cases with a larger time difference between the disease onset and the PCL assessment had higher PCL total score (MVP: Mann-Kendall Trend Test tau = 0.932, p<2×10^−16^; UKB: Mann-Kendall Trend Test tau = 0.376, p=0.005; Figure 3). Stratifying CAD cases depending on whether the disease onset was before or after the PCL assessment, this relationship appears to be consistent with the possible negative mental health consequences associated with CAD and its symptoms (Table 1). Indeed, CAD cases with PCL assessment after their cardiac disease onset had higher PCL total score when the time difference between disease onset and PCL assessment was longer (MVP: beta=0.113, p=6.95×10^−16^; UKB: beta=0.011, p=0.027). Conversely, CAD cases with PCL assessment before cardiac disease onset had PCL-17 total score lower when the time difference between the disease diagnosis and PCL assessment was larger (MVP: beta=0.192, p=0.011).

**Table 1:** Association of CAD (status and time of the diagnosis) with PCL total scores in MVP and UKB. In the case-control analysis, the PCL total scores of controls were compared with those of CAD cases (total sample and stratified by the timing of their cardiac disease onset with respect to PCL assessment). In the case-only analysis, we tested the association of PCL total scores with the temporal difference (years) between the time of CAD diagnosis and the time of PCL assessment. In MVP, the analyses were corrected for age, sex, income, self-reported ethnicity, and self-reported racial background. In UKB, the analyses were corrected for age, sex, Townsend deprivation index, and self-reported ethnic background.

| CAD | MVP (PCL-17; control N=245,962) |  |  |  |  |  |  | UKB (PCL-6, control N=146,875) |  |  |  |  |  |  |
| --- | --- | --- | --- | --- | --- | --- | --- | --- | --- | --- | --- | --- | --- | --- |
|  | case<br>N | Case-Control |  |  | Case-only |  |  | case<br>N | Case-Control |  |  | Case-only |  |  |
|  |  | Beta | SE | P | Beta | SE | P |  | Beta | SE | P | Beta | SE | P |
| **Total sample** | 73,074 | 2.89 | 0.06 | $<10^{-300}$ | 0.112 | 0.01 | $4.08 \times 10^{-29}$ | 8,942 | 0.584 | 0.034 | $3.98 \times 10^{-66}$ | 0.013 | 0.004 | 0.001 |
| **Onset after PCL assessment** | 12,267 | 2.19 | 0.13 | $1.12 \times 10^{-63}$ | 0.192 | 0.075 | 0.011 | 167 | 0.248 | 0.234 | 0.391 | -0.567 | 0.533 | 0.287 |
| **Onset same-year PCL assessment** | 4,646 | 2.71 | 0.21 | $4.23 \times 10^{-38}$ | - | - | - | 590 | 0.443 | 0.125 | $3.94 \times 10^{-4}$ | - | - | - |
| **Onset before PCL assessment** | 56,161 | 3.027 | 0.07 | $<10^{-300}$ | 0.113 | 0.014 | $6.95 \times 10^{-16}$ | 8,185 | 0.6 | 0.035 | $7.11 \times 10^{-66}$ | 0.011 | 0.005 | 0.027 |

**Figure 3:**
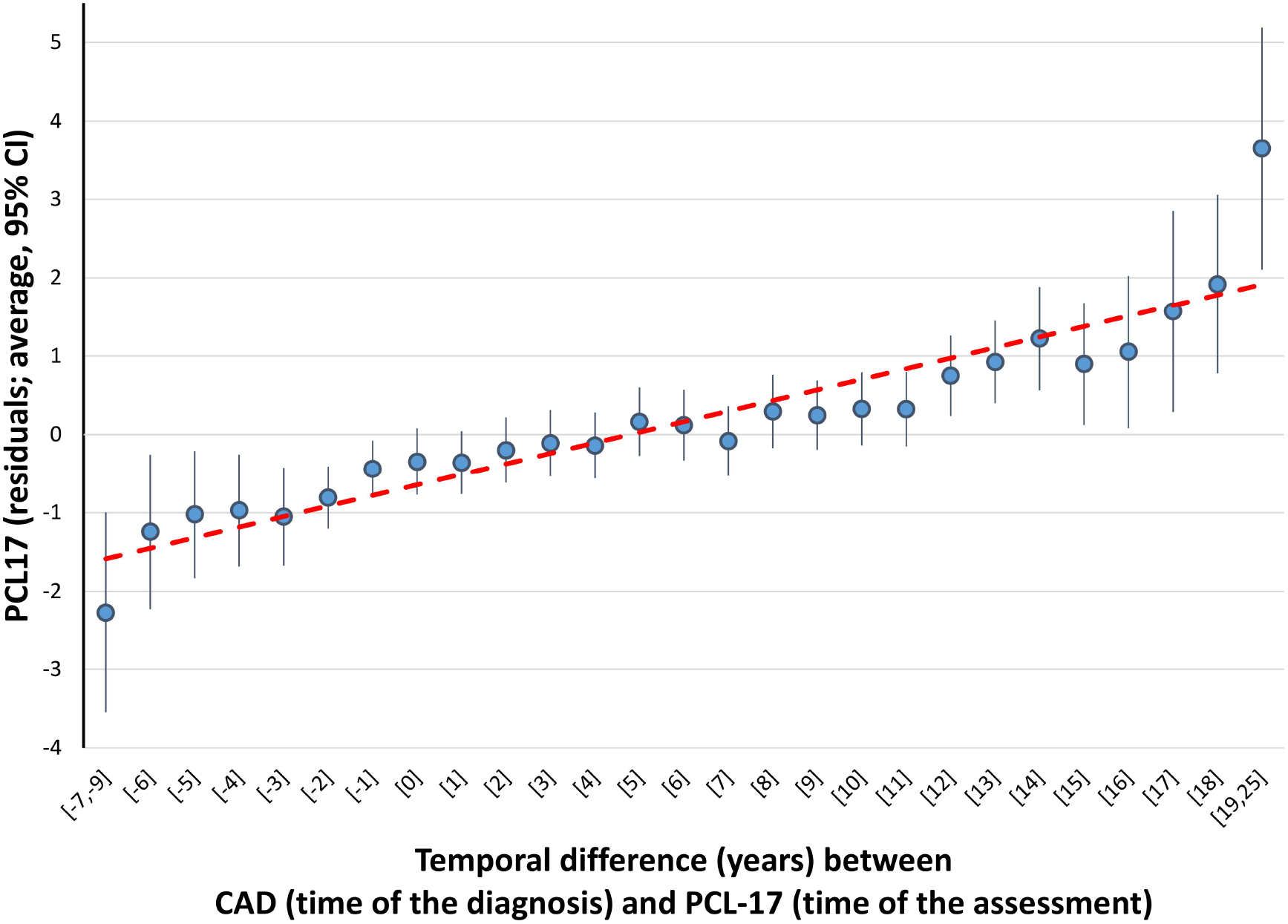
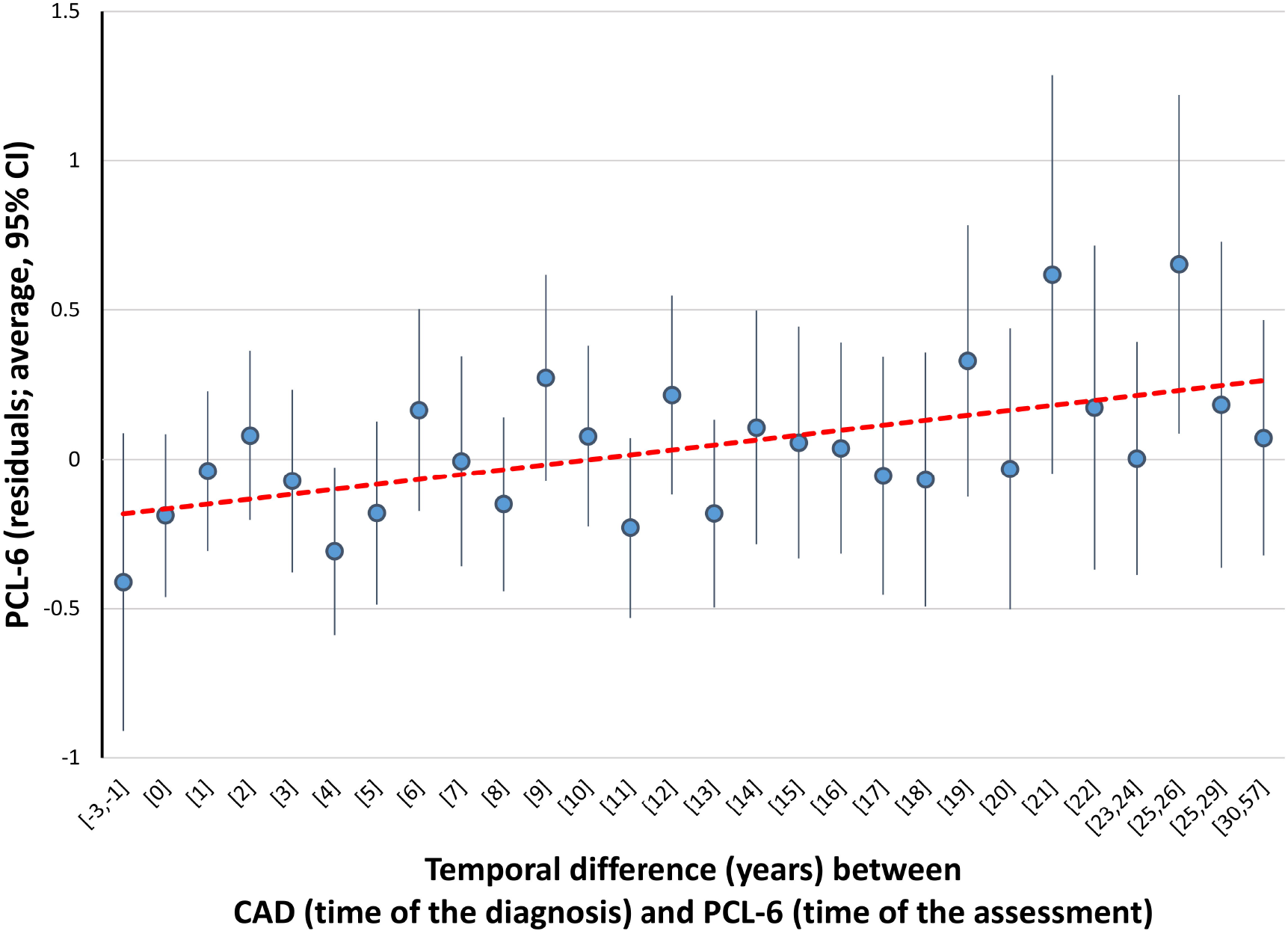
Relationship of PCL total scores with the temporal difference between CAD (time of the diagnosis) and PCL (time of the assessment) in MVP (**A**) and UKB (**B**). In MVP, PCL-17 residuals were obtained regressing PCL-17 total score on age, sex, income, self-reported ethnicity, and self-reported racial background. In UKB, PCL-6 residuals were obtained regressing PCL-6 total score on age, sex, Townsend deprivation index, and self-reported ethnic background.

## DISCUSSION

Understanding the dynamics underlying the comorbidities of posttraumatic stress and CAD can have major implications to improve our ability to provide more comprehensive patient care. In the present study, we used large-scale datasets including GWA statistics and EHRs to explore whether cause-effect relationships are responsible for the comorbidity observed between PTSD and CAD.

Leveraging GWA statistics generated by consortium meta-analyses and biobanks, we assessed the genetic correlation of CAD with PTSD, defining PTSD via case-control or quantitative measures. We observed consistent results across datasets converging on ∼20% CAD-PTSD genetic correlation. Conversely, when testing traits related to traumatic experience and social support, there was a lack of consistent pleiotropy with CAD across the datasets investigated. This supports the notion that the PTSD-CAD comorbidity can be partially due to pleiotropic mechanisms by which some risk loci contribute to both diseases, but these shared genetic effects did not appear to be due to the association of PTSD with traumatic events and social support. The only exception was the shared genetic correlation (rg=∼0.5) of PTSD and CAD with “Been diagnosed with life-threatening disease”. This likely reflects CAD prevalence among life-threatening diseases^57^. CAD diagnosis itself and the medical encounters during which it is diagnosed may be experienced as traumatic experiences leading to posttraumatic stress^58^.

To disentangle the possible mechanisms underlying the PTSD-CAD genetic correlation, we conducted a genetically-informed causal inference analysis. In line with previous findings^28^, we observed that PCL-17 total score is statistically more powerful than the other PTSD-related traits. Since traits derived from PCL-17 are highly genetically correlated with PTSD case-control definition (rg>0.95; Supplemental Table 1), we considered PCL-17 total score as the primary phenotype for our two-sample MR analysis with CAD. We observed a significant bidirectional relationship between CAD and PCL-17 total score. The effect of genetically-determined PCL-17 total score on CAD was in line with the positive genetic correlation observed between these two traits: an increase of one standard deviation in the PCL-17 total score was associated with a 3.5% increase in the odds for CAD risk. This putative causal effect could contribute to the observed association of PTSD with CVD outcomes reported in epidemiological studies^4^.

Conversely, we observed an effect of genetically-determined CAD on PCL-17 total score where a unit increase in the log(Odds Ratio) for CAD was associated with a 0.4 *decrease* in the PCL-17 total score. This appears to contradict epidemiological evidence^4^ and the overall PTSD-CAD genetic correlation we estimated across multiple datasets. To verify this unexpected finding, we investigated possible biases due to heterogeneity and horizontal pleiotropy within the instrumental variable and the reliability of the estimates across MR methods, data sources, and PTSD outcomes. The results are consistent across these sensitivity analyses. However, testing traits derived from PCL-6 as exposure, we observed a null effect with respect to CAD. Unfortunately, no large-scale GWAS of PCL-17-derived traits is available other than the one conducted in MVP, so we cannot exclude that the inverse effect of CAD on PCL-17-derived traits is due to the particular structure of the MVP cohort (or differences in patients within the VA healthcare system). Alternatively, the observed negative effect may be also due to a survival bias in CAD genetic associations although a previous study indicated this should be very limited^59^.

Our pleiotropic meta-analysis to investigate loci with concordant and discordant effects between PCL-17 and CAD highlighted different biological processes. Loci with concordant effect showed enrichment for platelet amyloid precursor protein pathway and negative regulation of astrocyte activation. APP is a major inhibitor of platelet aggregation and is released from activated platelets in patients with acute coronary syndrome^60,61^. APP demonstrated neuroprotective properties following traumatic brain injuries, including long-term neuronal survival and neuronal extension^62^. In this context, the regulation of astrocyte activation suggests that loci with concordant effects between PCL17 and CAD are involved in pathways related to neuroprotective functions. These may contribute to the comorbidity of trauma and PTSD with dementias^63,64^. Conversely, the loci with discordant effects between PCL-17 and CAD were enriched for multiple pathways related to lipid metabolism. Increased lipid levels (triglycerides and LDL) are associated with adverse cardiac outcomes^65^. With respect to PTSD, altered lipid levels are observed among affected patients in the context of interrelated somatic features including a proinflammatory milieu, metabolomic changes, and metabolic dysregulation^66^. This complex interplay may contribute to the negative effect of CAD genetic liability on PCL-17.

We used EHRs from MVP and UKB to investigate the longitudinal changes in the association between CAD and PTSD symptom severity. Specifically, we tested how the time difference between the PCL assessment and the CAD diagnosis affects the relationship of CAD with PTSD symptom severity. For the case-control comparisons, CAD is associated with increased PCL total score in both MVP and UKB cohorts, including when stratifying cases by whether the CAD diagnosis was received before, after, or in the same year as the PCL assessment. We also conducted a case-only analysis investigating whether there is a relationship between the time difference of PCL assessment and CAD diagnosis. Among CAD cases diagnosed before the PCL assessment, an earlier CAD diagnosis was associated with a higher PCL total score later in life in both MVP and UKB cohorts. This is in line with the fact CAD is a life-threatening disease that strongly impacts the quality of life of the individuals affected. A growing literature regarding cardiac-disease-induced PTSD highlights the negative mental outcomes associated with acute coronary syndrome and related medical procedures^67^. Additionally, the enduring somatic threat model has been proposed to address PTSD due to acute manifestations of chronic diseases that are enduring/internal in nature^58^. However, these reports suggest that the onset of CAD is followed closely in time with PTSD. Conversely, our EHR-based findings highlight the potential long-term effect of CAD on PTSD severity.

With respect to CAD subjects who received their CAD diagnosis after the PCL assessment, MVP participants with lower PCL-17 total score had received a CAD diagnosis later in time than individuals with higher PCL-17 total score. This association is in line with the positive association of genetically-determined PCL-17 on CAD identified in our two-sample MR analysis. However, we cannot exclude the possibility that individuals receiving a CAD diagnosis closer to the time of their PCL assessment may at the time be experiencing cardiac symptoms that increase their psychological stress and PTSD symptoms.

In line with MR (bidirectional relationship) and LCV (no significant genetic causal proportion), our EHR-based follow-up analysis did not find direct evidence supporting a causal effect between CAD and PTSD severity. However, we found the relationship between CAD and PTSD severity is affected by the timing of CAD onset. A recent study showed that, although earlier health-event occurrence is genetically correlated with a higher polygenic risk of disease susceptibility, age of first occurrence has specific genetic risk factors not associated with susceptibility to the disease^68^. With respect to outcomes related to the circulatory system, the genetic correlation between age of first occurrence and disease susceptibility was -0.56^68^, highlighting the presence of an independent genetic component between them. Because of the effect of CAD onset on PTSD severity, we speculate that the association of higher CAD genetic liability with lower PCL-17 score could be due to the unaccounted effect of the genetic predisposition to cardiac disease onset among CAD cases. Unfortunately, there is no large-scale GWAS of CAD onset to test this hypothesis using genetically-informed causal inference methods.

Although the present study provides novel insights into the complex relationship between PTSD and CAD, we acknowledge several limitations. Genetically-informed causal inference analysis was conducted using genome-wide association statistics generated from cohorts including individuals of European descent because of the unavailability of large-scale PTSD and CAD GWAS in other ancestry groups. In the EHR-based follow-up analysis, we investigate UKB and MVP participants of different racial and ethnic backgrounds considering the latter as a covariate in the regression analysis. Due to the limited sample size available, we did not explore possible differences across racial and ethnic backgrounds. Accordingly, further studies will be needed to investigate genetic and phenotype differences in PTSD-CAD relationships across worldwide populations. Because of the higher statistical power of the PCL-17 GWAS, our primary analysis tested this cross-sectional assessment with respect to CAD (a lifetime diagnosis). Although PCL-17 can surely vary across time, the high genetic correlation between PTSD and PCL-17 (rg>0.95) strongly supports that PCL-17 cross-sectional assessment is a very good proxy of PTSD lifetime diagnosis. Finally, we did not explore the effect of possible mediators in the PTSD-CAD relationship, because our main goal was to test the bidirectional associations between PTSD and CAD across different datasets and methods. Further studies will be needed to investigate whether the effects detected are due to risk factors shared between PTSD and CAD.

In conclusion, our findings highlight the complex relationship between PTSD severity and CAD. Both our genetically-informed causal inference analysis and our EHR-based follow-up investigation highlighted a bidirectional relationship between PTSD and CAD. This strongly highlights the need for a more integrated approach toward the study of comorbidities of outcomes related to mental and physical health.

## Data Availability

All data produced in the present work are contained in the manuscript

## CONTRIBUTORS

RP, CJO, TLA, MBS, JG designed the study. RP, FRW, GAP, DST, CT, ATH, DFL, KA, JMG, CJO, TLA, MBS, JG collected and interpreted the data. RP, FRW, GAP, DST analyzed the data. RP wrote the manuscript. RP, FRW, GAP, DST, CT, ATH, DFL, KA, JMG, CJO, TLA, MBS, JG edited the manuscript. RP, CJO, TLA, MBS, JG supervised the study.

## DATA SHARING

All data used discussed in this study are provided as Supplementary Material.

## ACKNOWLEDGEMENTS

This research is based on data from the MVP, Office of Research and Development, Veterans Health Administration and was supported by funding from the VA Cooperative Studies Program (CSP, no. CSP575B) and the Veterans Affairs Office of Research and Development MVP (grant nos. MVP000 and VA Merit MVP025). The views expressed in this article are those of the authors and do not necessarily reflect the position or policy of the Department of Veterans Affairs or the United States government. We thank the veterans who participated in this study, and the members of the VA CSP and MVP study teams, without whom this work would not have been possible. This research has been also conducted using the UK Biobank Resource (application reference no. 58146).

