## Supplemental Figures for "Understanding the comorbidity between posttraumatic stress severity and coronary artery disease using genome-wide information and electronic health records"

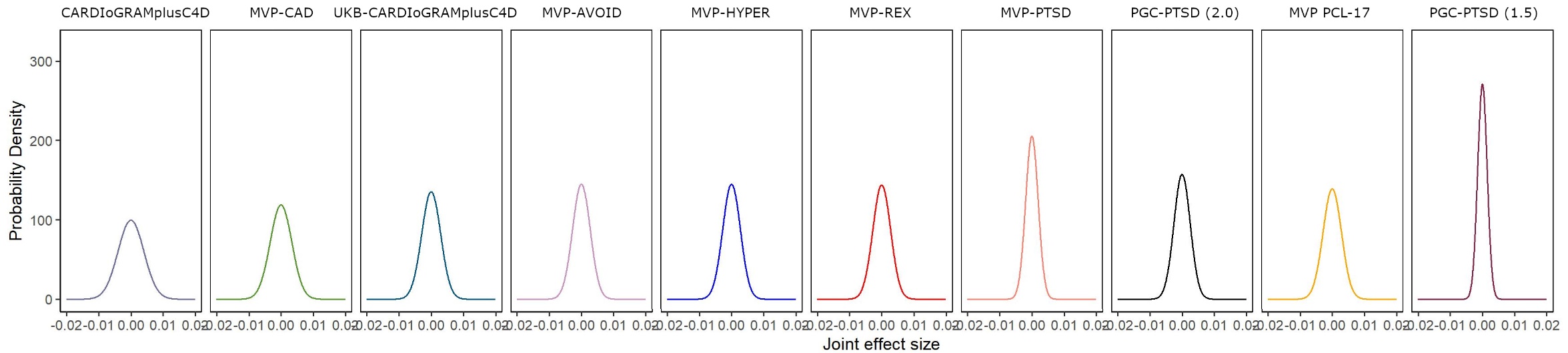

**Supplemental Figure 1**: Effect size distribution curves of the CAD and PTSD traits investigated.

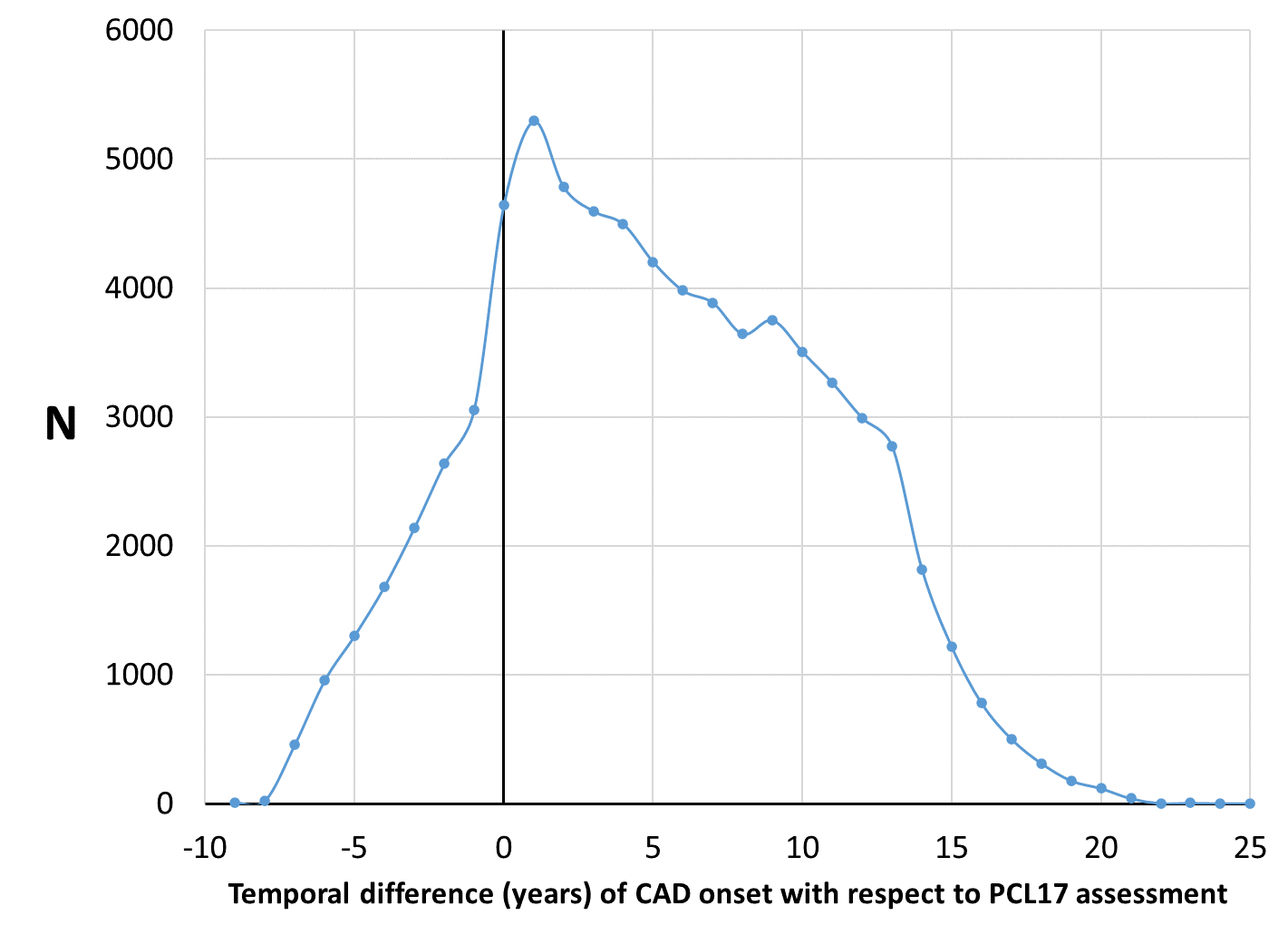

**Supplemental Figure 2**: Sample size distribution of MVP CAD cases in relation to the temporal difference (year) of the disease onset with respect to the time of the PCL-17 assessment.

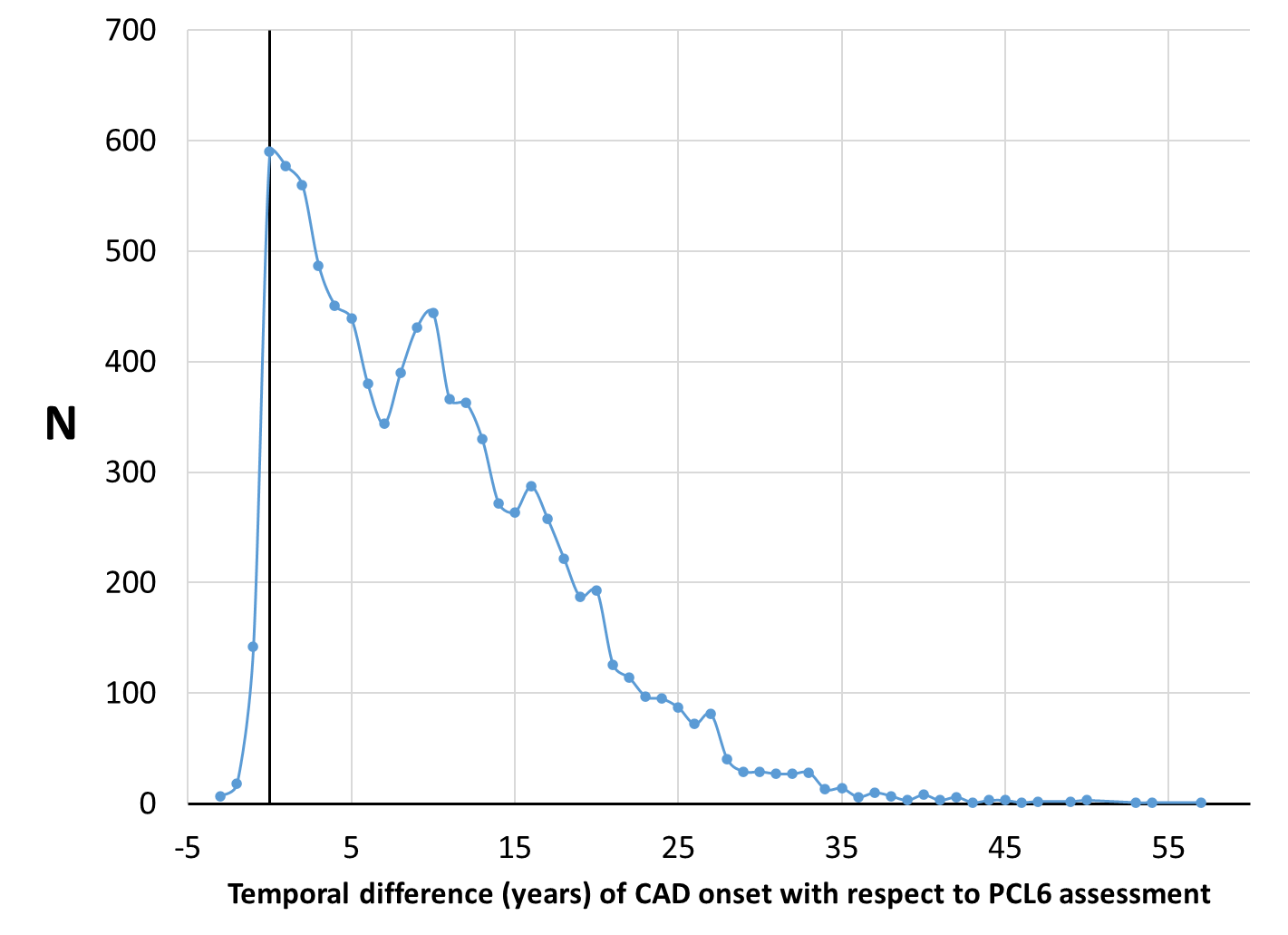

**Supplemental Table 3**: Sample size distribution of UKB CAD cases in relation to the temporal difference (year) of the disease onset with respect to the time of the PCL-6 assessment.
