## Supplemental Tables for "Understanding the comorbidity between posttraumatic stress severity and coronary artery disease using genome-wide information and electronic health records"

**Supplemental Table 1: Genetic correlation of PTSD (A) and CAD (B) among different datasets.**

| (A) | MVP PTSD | | | | | | |
|---|---|---|---|---|---|---|---|
| PGC-PTSD v2 | 0.961 (p=2.48e-17) | PGC-PTSD v2 | | | | | |
| PGC-PTSD v1.5 | 1 (p=1.45e-5) | 1 (p=3.91e-35) | PGC-PTSD v1.5 | | | | |
| MVP PCL17 | 0.961 (p=3.99e-247) | 0.924 (p=2.20e-21) | 0.953 (p=2.42e-5) | MVP PCL17 | | | |
| MVP REX | 0.977 (p=5.17e-238) | 0.890 (p=5.23e-22) | 0.930 (p=1.88e-6) | 0.973 (p<1e-300) | MVP REX | | |
| MVP AVOID | 0.915 (p=1.04e-201) | 0.922 (p=5.92e-22) | 0.916 (p=8.51e-6) | 0.984 (p<1e-300) | 0.931 (p=1e-300) | MVP AVOID | |
| MVP HYPER | 0.953 (p=3.52e-239) | 0.877 (p=2.77e-23) | 0.835 (p=1.51e-05) | 0.979 (p<1-300) | 0.935 (p=1e-300) | 0.943 (p=1e-300) | MVP HYPER |

| (B) | MVP CAD | | |
|---|---|---|---|
| UKB.CARDIoGRAMplusC4D CAD | 0.973 (p=1.01e-299) | UKB.CARDIoGRAMplusC4D CAD | |
| CARDIoGRAMplusC4D CAD | 1 (p=2.4e-244) | 1 (p<1e-300) | CARDIoGRAMplusC4D CAD |

**Supplemental Table 2: Genetic correlation between PTSD and CAD traits.**

| | MVP-CAD | UKB-CARDIoGRAMplusC4D-CAD | CARDIoGRAMplusC4D-CAD |
|---|---|---|---|
| **MVP-PTSD** | 0.181 (p=1.25e-5) | 0.221 (p=8.768e-9) | 0.201 (p=1.51e-6) |
| **MVP-PCL17** | 0.195 (p=6.71e-9) | 0.216 (p=7.43e-11) | 0.198 (p=4.09e-6) |
| **MVP-PCL6** | 0.154 (p=2.85e-6) | 0.202 (p=9.2e-13) | 0.163 (p=1.87e-7) |
| **MVP-AVO** | 0.208 (p=5.54e-10) | 0.211 (p=1.39e-10) | 0.191 (p=2.94e-8) |
| **MVP-REX** | 0.188 (p=9.36e-9) | 0.22 (p=1.36e-13) | 0.199 (p=9.13e-11) |
| **MVP-HYP** | 0.217 (p=2.67e-12) | 0.216 (p=5.96e-12) | 0.185 (p=7.17e-8) |
| **PGC-PTSD2** | 0.207 (p=9e-4) | 0.26 (p=1.76e-5) | 0.255 (p=3e-4) |
| **PGC-PTSD1-5** | 0.195 (p=0.035) | 0.299 (p=0.004) | 0.324 (p=0.007) |

**Supplemental Table 3: Genetic correlation of CAD with traits related to traumatic events and social support.**

| UKB Field ID - Description | MVP-CAD | UKB-CARDIoGRAMplusC4D-CAD | CARDIoGRAMplusC4D-CAD |
|---|---|---|---|
| 20489 Felt loved as a child | -0.005 (p=0.873) | -0.096 (p=0.009) | -0.063 (p=0.140) |
| 20488 Physically abused by family as a child | 0.047 (p=0.295) | 0.148 (p=0.001) | 0.088 (p=0.099) |
| 20487 Felt hated by family member as a child | 0.069 (p=0.102) | 0.171 (p=7.72e-5) | 0.122 (p=0.021) |
| 20490 Sexually molested as a child | 0.058 (p=0.258) | 0.213 (p=3e-4) | 0.174 (p=0.012) |
| 20491 Someone to take to doctor when needed as a child | 0.003 (p=0.955) | -0.205 (p=2e-4) | -0.143 (p=0.012) |
| 20522 Been in a confiding relationship as an adult | 0.019 (p=0.685) | -0.0271 (p=0.517) | -0.006 (p=0.907) |
| 20523 Physical violence by partner or ex-partner as an adult | 0.018 (p=0.752) | 0.162 (p=0.001) | 0.101 (p=0.052) |
| 20521 Belittlement by partner or ex-partner as an adult | -0.01 (p=0.866) | 0.135 (p=0.004) | 0.097 (p=0.058) |
| 20524 Sexual interference by partner or ex-partner without consent as an adult | 0.007 (p=0.921) | 0.178 (p=0.013) | 0.126 (p=0.121) |
| 20525 Able to pay rent/mortgage as an adult | -0.09 (p=0.152) | -0.237 (p=0.0002) | -0.175 (p=0.01) |
| 20531 Victim of sexual assault | -0.056 (p=0.251) | 0.05 (p=0.331) | 0.035 (p=0.535) |
| 20529 Victim of physically violent crime | -0.13 (p=0.019) | -0.022 (p=0.706) | -0.033 (p=0.625) |
| 20526 Been in serious accident believed to be life-threatening | 0.015 (p=0.827) | 0.152 (p=0.044) | 0.153 (p=0.047) |
| 20530 Witnessed sudden violent death | 0.039 (p=0.525) | 0.16 (p=0.013) | 0.142 (p=0.037) |
| 20528 Diagnosed with life-threatening illness | 0.338 (p=5.78e-5) | 0.53 (p=2.89e-7) | 0.528 (p=9.37e-7) |
| 20527 Been involved in combat or exposed to war-zone | 0.137 (p=0.059) | 0.101 (p=0.215) | 0.149 (p=0.113) |

**Supplemental Table 4: Genetic correlaiton of "Diagnosed with life-threatening illness" with PTSD and CAD traits.**

| | **20528_Diagnosed with life-threatening illness** |
|---|---|
| **MVP-PTSD** | 0.294 (p=0.009) |
| **MVP-PCL17** | 0.495 (p=1.28e-6) |
| **MVP-AVO** | 0.508 (p=1.02e-6) |
| **MVP-REX** | 0.437 (p=7.62e-6) |
| **MVP-HYP** | 0.452 (p=4.77e-6) |
| **PGC-PTSD2** | 0.645 (p=2e-4) |
| **PGC-PTSD1-5** | 0.525 (p=0.034) |
| **MVP-CAD** | 0.338 (p=5.78e-5) |
| **UKB-CARDIoGRAMplusC4D-CAD** | 0.53 (p=2.89e-7) |
| **CARDIoGRAMplusC4D-CAD** | 0.528 (p=9.37e-7) |

**Supplemental Table 5: PRS analysis (threshold P<5E0-8) between PTSD and CAD traits.**

| Training | Target | r2 | P |
|---|---|---|---|
| MVP-AVOI | UKB-CARDIO-CAD | 2.66E-05 | 0.020 |
| MVP-HYPE | UKB-CARDIO-CAD | 9.93E-05 | 3.35E-05 |
| MVP-PCL | UKB-CARDIO-CAD | 1.11E-04 | 1.30E-05 |
| MVP-PTSD | UKB-CARDIO-CAD | 3.79E-05 | 0.007 |
| MVP-REX | UKB-CARDIO-CAD | 1.45E-05 | 0.064 |
| UKB-CARDIO-CAD | MVP-AVOI | 5.31E-05 | 0.002 |
| UKB-CARDIO-CAD | MVP-HYPE | 6.31E-05 | 6.32E-04 |
| UKB-CARDIO-CAD | MVP-PCL | 6.75E-05 | 4.29E-04 |
| UKB-CARDIO-CAD | MVP-PTSD | 4.21E-05 | 0.004 |
| UKB-CARDIO-CAD | MVP-REX | 6.01E-05 | 8.32E-04 |

**Supplemental Table 6: bidirectional MR analyses between PCL-17 (MVP) on CAD (UKB-CARDIoGRAMplusC4D).**

| PCL-17 (MVP) → CAD (UKB-CARDIoGRAMplusC4D) | | | | |
|---|---|---|---|---|
| **MR method** | **nsnp** | **b** | **se** | **pval** |
| **MR Egger** | 13 | 0.08 | 0.07 | 0.322 |
| **Weighted median** | 13 | 0.04 | 0.01 | 0.002 |
| **Inverse variance weighted** | 13 | 0.03 | 0.01 | 0.003 |
| **Simple mode** | 13 | 0.04 | 0.02 | 0.095 |
| **Weighted mode** | 13 | 0.04 | 0.02 | 0.063 |
| | **Estimate** | **se** | **pval** | |
| **MR Egger - Intercept** | -0.014 | 0.024 | 0.572 | |
| | **Q** | **Q_df** | **Q_pval** | |
| **Maximum likelihood - Heterogeneity test** | 22.5 | 12 | 0.032 | |
| **MR Egger - Heterogeneity test** | 22.6 | 11 | 0.020 | |
| **Inverse variance weighted - Heterogeneity test** | 23.3 | 12 | 0.025 | |
| | **Estimate** | **SE** | **P** | |
| **MR-RAPS - Causal effect** | 0.035 | 0.012 | 0.004 | |
| **MR-RAPS - Pleiotropy variance** | 8.90E-05 | 7.55E-05 | 0.238 | |
| | **RSSobs** | **Pvalue** | | |
| **MR-PRESSO Global Test** | 26.9 | 0.037 | | |

| CAD (UKB-CARDIoGRAMplusC4D) → PCL-17 (MVP) | | | | |
|---|---|---|---|---|
| **MR method** | **nsnp** | **b** | **se** | **pval** |
| **MR Egger** | 33 | -0.82 | 0.45 | 0.079 |
| **Weighted median** | 33 | -0.45 | 0.18 | 0.011 |
| **Inverse variance weighted** | 33 | -0.43 | 0.19 | 0.029 |
| **Simple mode** | 33 | -0.36 | 0.32 | 0.265 |
| **Weighted mode** | 33 | -0.55 | 0.20 | 0.011 |
| | **Estimate** | **se** | **pval** | |
| **MR Egger - Intercept** | 0.038 | 0.039 | 0.341 | |
| | **Q** | **Q_df** | **Q_pval** | |
| **Maximum likelihood - Heterogeneity test** | 89.1 | 32 | 2.72E-07 | |
| **MR Egger - Heterogeneity test** | 86.8 | 31 | 3.43E-07 | |
| **Inverse variance weighted - Heterogeneity test** | 89.4 | 32 | 2.45E-07 | |
| | **Estimate** | **SE** | **P** | |
| **MR-RAPS - Causal effect** | -0.407 | 0.169 | 0.016 | |
| **MR-RAPS - Pleiotropy variance** | | | | |
| | **RSSobs** | **Pvalue** | | |
| **MR-PRESSO Global Test** | 110.9 | <1e-04 | | |

**Supplemental Table 7: Leave-one-out (LOO) analysis assessing the heterogeneity within the genetic instrument testing CAD (UKB-CARDIoGRAMplusC4D) → PCL-17 (MVP). Outelier variants are highlighted in red.**

| rsid | -log10(P value - IVW Heterogeneity test) | Z score (LOO - IVW Heteorgeneity test) |
|---|---|---|
| rs7623687 | 4.36 | -3.35 |
| rs2071382 | 4.45 | -3.21 |
| rs11191416 | 5.38 | -1.72 |
| rs4299376 | 5.64 | -1.29 |
| rs2244608 | 6.09 | -0.57 |
| rs585967 | 6.27 | -0.28 |
| rs10774625 | 6.40 | -0.08 |
| rs2681472 | 6.42 | -0.04 |
| rs12202017 | 6.45 | 0.01 |
| rs6511720 | 6.45 | 0.01 |
| rs2891168 | 6.48 | 0.05 |
| rs62265630 | 6.60 | 0.24 |
| rs7164479 | 6.61 | 0.26 |
| rs6841581 | 6.61 | 0.26 |
| rs3918226 | 6.62 | 0.27 |
| rs72743461 | 6.64 | 0.31 |
| rs16986953 | 6.69 | 0.38 |
| rs9349379 | 6.73 | 0.44 |
| rs6909752 | 6.73 | 0.45 |
| rs2306556 | 6.73 | 0.45 |
| rs1887318 | 6.74 | 0.46 |
| rs10840293 | 6.74 | 0.47 |
| rs2107595 | 6.76 | 0.51 |
| rs11556924 | 6.78 | 0.54 |
| rs507666 | 6.78 | 0.54 |
| rs10080815 | 6.79 | 0.54 |
| rs2246942 | 6.81 | 0.58 |
| rs28451064 | 6.83 | 0.61 |
| rs10455872 | 6.84 | 0.62 |
| rs1870634 | 6.84 | 0.63 |
| rs7500448 | 6.84 | 0.63 |
| rs7412 | 6.85 | 0.64 |
| rs1250229 | 6.85 | 0.64 |

**Supplemental Table 8: MR analysis testing the effect of CAD (UKB-CARDIoGRAMplusC4D) on PCL-17 (MVP) after removal of outlier variants from the instrumental.variable**

| MR method | nsnp | b | se | pval |
|---|---|---|---|---|
| MR Egger | 29 | -0.78 | 0.31 | 0.019 |
| Weighted median | 29 | -0.50 | 0.18 | 0.005 |
| Inverse variance weighted | 29 | -0.50 | 0.14 | 2.61E-04 |
| Simple mode | 29 | -0.26 | 0.36 | 0.467 |
| Weighted mode | 29 | -0.56 | 0.21 | 0.012 |

| | Estimate | se | pval |
|---|---|---|---|
| MR Egger - Intercept | 0.028 | 0.028 | 0.335 |

| | Q | Q_df | Q_pval |
|---|---|---|---|
| Maximum likelihood - Heterogeneity test | 36.7 | 28 | 0.125 |
| MR Egger - Heterogeneity test | 35.6 | 27 | 0.124 |
| Inverse variance weighted - Heterogeneity test | 36.9 | 28 | 0.121 |

| | Estimate | SE | P |
|---|---|---|---|
| MR-RAPS - Causal effect | -0.52 | 0.151 | 5.47E-04 |
| MR-RAPS - Pleiotropy variance | 1.56E-03 | 1.38E-03 | 0.256 |

| | RSSobs | Pvalue |
|---|---|---|
| MR-PRESSO Global Test | 39.4 | 0.137 |

**Supplemental Table 9: MR analysis testing the association between CAD (UKB-CARDIoGRAMplusC4D) and PTSD symptoms (MVP HYPE, REX, and AVOID).**

**CAD (UKB-CARDIoGRAMplusC4D) → HYPE (MVP)**

| MR method | nsnp | b | se | pval |
|---|---|---|---|---|
| MR Egger | 39 | -0.25 | 0.13 | 0.066 |
| Weighted median | 39 | -0.13 | 0.05 | 0.019 |
| Inverse variance weighted | 39 | -0.12 | 0.06 | 0.030 |
| Simple mode | 39 | -0.11 | 0.10 | 0.293 |
| Weighted mode | 39 | -0.16 | 0.06 | 0.010 |
| | **Estimate** | **se** | **pval** | |
| MR Egger - Intercept | 0.012 | 0.011 | 0.296 | |
| | **Q** | **Q_df** | **Q_pval** | |
| Maximum likelihood - Heterogeneity test | 98.5 | 38 | 2.88E-07 | |
| MR Egger - Heterogeneity test | 95.9 | 37 | 4.05E-07 | |
| Inverse variance weighted - Heterogeneity test | 98.8 | 38 | 2.62E-07 | |
| | **Estimate** | **SE** | **P** | |
| MR-RAPS - Causal effect | -0.117 | 0.049 | 0.018 | |
| MR-RAPS - Pleiotropy variance | 4.42E-04 | 1.77E-04 | 0.012 | |
| | **RSSobs** | **Pvalue** | | |
| MR-PRESSO Global Test | 115.5 | <1e-04 | | |

**HYPE (MVP) → CAD (UKB-CARDIoGRAMplusC4D)**

| MR method | nsnp | b | se | pval |
|---|---|---|---|---|
| MR Egger | 9 | 0.21 | 0.23 | 0.399 |
| Weighted median | 9 | 0.07 | 0.04 | 0.096 |
| Inverse variance weighted | 9 | 0.09 | 0.04 | 0.008 |
| Simple mode | 9 | 0.02 | 0.07 | 0.783 |
| Weighted mode | 9 | 0.02 | 0.07 | 0.782 |
| | **Estimate** | **se** | **pval** | |
| MR Egger - Intercept | -0.012 | 0.024 | 0.634 | |
| | **Q** | **Q_df** | **Q_pval** | |
| Maximum likelihood - Heterogeneity test | 11.1 | 8 | 0.199 | |
| MR Egger - Heterogeneity test | 11.0 | 7 | 0.139 | |
| Inverse variance weighted - Heterogeneity test | 11.4 | 8 | 0.181 | |
| | **Estimate** | **SE** | **P** | |
| MR-RAPS - Causal effect | 0.090 | 0.028 | 0.001 | |
| MR-RAPS - Pleiotropy variance | 0 | 0 | NaN | |
| | **RSSobs** | **Pvalue** | | |
| MR-PRESSO Global Test | 13.7 | 0.422 | | |

**CAD (UKB-CARDIoGRAMplusC4D) → REX (MVP)**

| MR method | nsnp | b | se | pval |
|---|---|---|---|---|
| MR Egger | 39 | -0.22 | 0.12 | 0.074 |
| Weighted median | 39 | -0.11 | 0.05 | 0.040 |
| Inverse variance weighted | 39 | -0.13 | 0.05 | 0.015 |
| Simple mode | 39 | -0.05 | 0.10 | 0.608 |
| Weighted mode | 39 | -0.10 | 0.06 | 0.127 |
| | **Estimate** | **se** | **pval** | |
| MR Egger - Intercept | 0.009 | 0.011 | 0.384 | |
| | **Q** | **Q_df** | **Q_pval** | |
| Maximum likelihood - Heterogeneity test | 86.7 | 38 | 1.15E-05 | |
| MR Egger - Heterogeneity test | 85.2 | 37 | 1.15E-05 | |
| Inverse variance weighted - Heterogeneity test | 87.0 | 38 | 1.05E-05 | |
| | **Estimate** | **SE** | **P** | |
| MR-RAPS - Causal effect | -0.122 | 0.045 | 0.006 | |
| MR-RAPS - Pleiotropy variance | 2.99E-04 | 1.40E-04 | 0.0325 | |
| | **RSSobs** | **Pvalue** | | |
| MR-PRESSO Global Test | 109.5 | <1e-04 | | |

**CAD (UKB-CARDIoGRAMplusC4D) → AVOI (MVP)**

| MR method | nsnp | b | se | pval |
|---|---|---|---|---|
| MR Egger | 39 | -0.32 | 0.16 | 0.057 |
| Weighted median | 39 | -0.15 | 0.08 | 0.045 |
| Inverse variance weighted | 39 | -0.15 | 0.07 | 0.037 |
| Simple mode | 39 | -0.23 | 0.15 | 0.127 |
| Weighted mode | 39 | -0.21 | 0.08 | 0.016 |
| | **Estimate** | **se** | **pval** | |
| MR Egger - Intercept | 0.017 | 0.014 | 0.246 | |
| | **Q** | **Q_df** | **Q_pval** | |
| Maximum likelihood - Heterogeneity test | 81.8 | 38 | 4.84E-05 | |
| MR Egger - Heterogeneity test | 79.0 | 37 | 7.11E-05 | |
| Inverse variance weighted - Heterogeneity test | 82.0 | 38 | 4.58E-05 | |
| | **Estimate** | **SE** | **P** | |
| MR-RAPS - Causal effect | -0.139 | 0.066 | 3.40E-02 | |
| MR-RAPS - Pleiotropy variance | 7.21E-04 | 3.08E-04 | 0.019 | |
| | **RSSobs** | **Pvalue** | | |
| MR-PRESSO Global Test | 102.3 | 1.00E-04 | | |

Supplemental Table 10: MR analysis testing the effect of CAD (UKB-CARDIoGRAMplusC4D) on PTSD symptoms (MVP HYPE, REX, and AVOID) after outlier variants removal from the instrumental variable.

| CAD (UKB-CARDIoGRAMplusC4D) → HYPE (MVP) | | | | |
|---|---|---|---|---|
| MR method | nsnp | b | se | pval |
| MR Egger | 34 | -0.25 | 0.09 | 0.011 |
| Weighted median | 34 | -0.13 | 0.06 | 0.019 |
| Inverse variance weighted | 34 | -0.14 | 0.04 | 4.72E-04 |
| Simple mode | 34 | -0.11 | 0.11 | 0.301 |
| Weighted mode | 34 | -0.17 | 0.06 | 0.014 |
| | Estimate | se | pval | |
| MR Egger - Intercept | 0.011 | 0.008 | 0.207 | |
| | Q | Q_df | Q_pval | |
| Maximum likelihood - Heterogeneity test | 40.4 | 33 | 0.175 | |
| MR Egger - Heterogeneity test | 38.6 | 32 | 0.196 | |
| Inverse variance weighted - Heterogeneity test | 40.6 | 33 | 0.170 | |
| | Estimate | SE | P | |
| MR-RAPS - Causal effect | -0.145 | 0.045 | 0.001 | |
| MR-RAPS - Pleiotropy variance | 1.52E-04 | 1.31E-04 | 0.246 | |
| | RSSobs | Pvalue | | |
| MR-PRESSO Global Test | 43.1 | 0.174 | | |

| CAD (UKB-CARDIoGRAMplusC4D) → AVOI (MVP) | | | | |
|---|---|---|---|---|
| MR method | nsnp | b | se | pval |
| MR Egger | 34 | -0.31 | 0.12 | 0.014 |
| Weighted median | 34 | -0.16 | 0.08 | 0.032 |
| Inverse variance weighted | 34 | -0.16 | 0.05 | 0.003 |
| Simple mode | 34 | -0.19 | 0.16 | 0.231 |
| Weighted mode | 34 | -0.21 | 0.10 | 0.047 |
| | Estimate | se | pval | |
| MR Egger - Intercept | 0.016 | 0.011 | 0.164 | |
| | Q | Q_df | Q_pval | |
| Maximum likelihood - Heterogeneity test | 35.8 | 33 | 0.337 | |
| MR Egger - Heterogeneity test | 33.8 | 32 | 0.382 | |
| Inverse variance weighted - Heterogeneity test | 35.9 | 33 | 0.334 | |
| | Estimate | SE | P | |
| MR-RAPS - Causal effect | -0.16 | 0.058 | 0.006 | |
| MR-RAPS - Pleiotropy variance | 1.82E-04 | 2.22E-04 | 0.412 | |
| | RSSobs | Pvalue | | |
| MR-PRESSO Global Test | 38.5 | 0.327 | | |

| CAD (UKB-CARDIoGRAMplusC4D) → REX (MVP) | | | | |
|---|---|---|---|---|
| MR method | nsnp | b | se | pval |
| MR Egger | 35 | -0.20 | 0.08 | 0.017 |
| Weighted median | 35 | -0.11 | 0.05 | 0.027 |
| Inverse variance weighted | 35 | -0.15 | 0.04 | 1.46E-05 |
| Simple mode | 35 | -0.05 | 0.10 | 0.638 |
| Weighted mode | 35 | -0.09 | 0.08 | 0.246 |
| | Estimate | se | pval | |
| MR Egger - Intercept | 0.005 | 0.007 | 0.489 | |
| | Q | Q_df | Q_pval | |
| Maximum likelihood - Heterogeneity test | 26.5 | 34 | 0.818 | |
| MR Egger - Heterogeneity test | 26.1 | 33 | 0.797 | |
| Inverse variance weighted - Heterogeneity test | 26.6 | 34 | 0.813 | |
| | Estimate | SE | P | |
| MR-RAPS - Causal effect | -0.153 | 0.037 | 3.17E-05 | |
| MR-RAPS - Pleiotropy variance | 0 | 0 | NaN | |
| | RSSobs | Pvalue | | |
| MR-PRESSO Global Test | 28.3 | 0.814 | | |

Supplemental Table 11: MR analysis testing the effect of CAD (CARDIoGRAMplusC4D) on PTSD symptoms (MVP PCL-17, HYPE, REX, and AVOID).

| CAD (CARDIoGRAMplusC4D) → PCL-17 (MVP) | | | | |
|---|---|---|---|---|
| MR method | nsnp | b | se | pval |
| MR Egger | 33 | -0.64 | 0.34 | 0.067 |
| Weighted median | 33 | -0.56 | 0.17 | 0.001 |
| Inverse variance weighted | 33 | -0.44 | 0.15 | 0.002 |
| Simple mode | 33 | 0.03 | 0.36 | 0.924 |
| Weighted mode | 33 | -0.45 | 0.19 | 0.027 |
| | Estimate | se | pval | |
| MR Egger - Intercept | 0.021 | 0.033 | 0.521 | |
| | Q | Q_df | Q_pval | |
| Maximum likelihood - Heterogeneity test | 59.9 | 32 | 0.002 | |
| MR Egger - Heterogeneity test | 59.4 | 31 | 0.002 | |
| Inverse variance weighted - Heterogeneity test | 60.2 | 32 | 0.002 | |
| | Estimate | SE | P | |
| MR-RAPS - Causal effect | -0.465 | 0.148 | 0.001 | |
| MR-RAPS - Pleiotropy variance | 0.003 | 1.00E-03 | 0.068 | |
| | RSSobs | Pvalue | | |
| MR-PRESSO Global Test | 67.2 | 0.003 | | |

| CAD (CARDIoGRAMplusC4D) → HYPE (MVP) | | | | |
|---|---|---|---|---|
| MR method | nsnp | b | se | pval |
| MR Egger | 38 | -0.21 | 0.10 | 0.051 |
| Weighted median | 38 | -0.15 | 0.05 | 0.003 |
| Inverse variance weighted | 38 | -0.14 | 0.04 | 0.002 |
| Simple mode | 38 | -0.15 | 0.09 | 0.106 |
| Weighted mode | 38 | -0.16 | 0.06 | 0.009 |
| | Estimate | se | pval | |
| MR Egger - Intercept | 0.008 | 0.010 | 0.446 | |
| | Q | Q_df | Q_pval | |
| Maximum likelihood - Heterogeneity test | 67.9 | 37 | 0.001 | |
| MR Egger - Heterogeneity test | 67.2 | 36 | 0.001 | |
| Inverse variance weighted - Heterogeneity test | 68.3 | 37 | 0.001 | |
| | Estimate | SE | P | |
| MR-RAPS - Causal effect | -0.144 | 0.043 | 0.001 | |
| MR-RAPS - Pleiotropy variance | 2.52E-04 | 1.45E-04 | 0.083 | |
| | RSSobs | Pvalue | | |
| MR-PRESSO Global Test | 73.5 | 0.002 | | |

| CAD (CARDIoGRAMplusC4D) → REX (MVP) | | | | |
|---|---|---|---|---|
| MR method | nsnp | b | se | pval |
| MR Egger | 38 | -0.16 | 0.09 | 0.092 |
| Weighted median | 38 | -0.10 | 0.05 | 0.031 |
| Inverse variance weighted | 38 | -0.13 | 0.04 | 1.24E-03 |
| Simple mode | 38 | -0.05 | 0.09 | 0.558 |
| Weighted mode | 38 | -0.09 | 0.06 | 0.121 |
| | Estimate | se | pval | |
| MR Egger - Intercept | 0.0036946 | 0.0092523 | 0.6920144 | |
| | Q | Q_df | Q_pval | |
| Maximum likelihood - Heterogeneity test | 59.9 | 37 | 0.010 | |
| MR Egger - Heterogeneity test | 59.9 | 36 | 0.007 | |
| Inverse variance weighted - Heterogeneity test | 60.2 | 37 | 0.009 | |
| | Estimate | SE | P | |
| MR-RAPS - Causal effect | -0.139 | 0.039 | 4.64E-04 | |
| MR-RAPS - Pleiotropy variance | 1.74E-04 | 1.21E-04 | 0.153 | |
| | RSSobs | Pvalue | | |
| MR-PRESSO Global Test | 68.5 | 0.008 | | |

| CAD (CARDIoGRAMplusC4D) → AVOI (MVP) | | | | |
|---|---|---|---|---|
| MR method | nsnp | b | se | pval |
| MR Egger | 38 | -0.27 | 0.13 | 0.043 |
| Weighted median | 38 | -0.11 | 0.07 | 0.111 |
| Inverse variance weighted | 38 | -0.15 | 0.06 | 8.71E-03 |
| Simple mode | 38 | -0.15 | 0.15 | 0.326 |
| Weighted mode | 38 | -0.17 | 0.09 | 0.068 |
| | Estimate | se | pval | |
| MR Egger - Intercept | 0.014 | 0.013 | 0.291 | |
| | Q | Q_df | Q_pval | |
| Maximum likelihood - Heterogeneity test | 57.8 | 37 | 0.016 | |
| MR Egger - Heterogeneity test | 56.2 | 36 | 0.017 | |
| Inverse variance weighted - Heterogeneity test | 58.0 | 37 | 0.015 | |
| | Estimate | SE | P | |
| MR-RAPS - Causal effect | -0.144 | 0.059 | 0.015 | |
| MR-RAPS - Pleiotropy variance | 4.76E-04 | 2.76E-04 | 0.084 | |
| | RSSobs | Pvalue | | |
| MR-PRESSO Global Test | 64.4 | 0.018 | | |

Supplemental Table 12: MR analysis testing the effect of CAD (CARDIoGRAMplusC4D) on PTSD symptoms (MVP PCL, HYPE, REX, and AVOID) after outlier variants removal from the instrumental variable.

| CAD (CARDIoGRAMplusC4D) → PCL-17 (MVP) | | | | |
|---|---|---|---|---|
| MR method | nsnp | b | se | pval |
| MR Egger | 30 | -0.60 | 0.28 | 0.042 |
| Weighted median | 30 | -0.59 | 0.17 | 4.68E-04 |
| Inverse variance weighted | 30 | -0.49 | 0.12 | 6.57E-05 |
| Simple mode | 30 | 0.10 | 0.40 | 0.805 |
| Weighted mode | 30 | -0.43 | 0.21 | 0.053 |
| | Estimate | se | pval | |
| MR Egger - Intercept | 0.012 | 0.028 | 0.668 | |
| | Q | Q_df | Q_pval | |
| Maximum likelihood - Heterogeneity test | 37.0 | 29 | 0.145 | |
| MR Egger - Heterogeneity test | 37.1 | 28 | 0.117 | |
| Inverse variance weighted - Heterogeneity test | 37.3 | 29 | 0.138 | |
| | Estimate | SE | P | |
| MR-RAPS - Causal effect | -0.506 | 0.138 | 2.00E-04 | |
| MR-RAPS - Pleiotropy variance | 1.59E-03 | 0.001 | 0.257 | |
| | RSSobs | Pvalue | | |
| MR-PRESSO Global Test | 40.0 | 0.145 | | |

| CAD (CARDIoGRAMplusC4D) → HYPE (MVP) | | | | |
|---|---|---|---|---|
| MR method | nsnp | b | se | pval |
| MR Egger | 36 | -0.19 | 0.08 | 0.032 |
| Weighted median | 36 | -0.16 | 0.05 | 0.001 |
| Inverse variance weighted | 36 | -0.15 | 0.04 | 2.81E-05 |
| Simple mode | 36 | -0.16 | 0.10 | 0.117 |
| Weighted mode | 36 | -0.16 | 0.06 | 0.015 |
| | Estimate | se | pval | |
| MR Egger - Intercept | 0.004 | 0.008 | 0.631 | |
| | Q | Q_df | Q_pval | |
| Maximum likelihood - Heterogeneity test | 42.6 | 35 | 0.177 | |
| MR Egger - Heterogeneity test | 42.6 | 34 | 0.149 | |
| Inverse variance weighted - Heterogeneity test | 42.9 | 35 | 0.170 | |
| | Estimate | SE | P | |
| MR-RAPS - Causal effect | -0.157 | 0.040 | 1.00E-04 | |
| MR-RAPS - Pleiotropy variance | 1.40E-04 | 1.28E-04 | 0.271 | |
| | RSSobs | Pvalue | | |
| MR-PRESSO Global Test | 45.2 | 0.184 | | |

| CAD (CARDIoGRAMplusC4D) → REX (MVP) | | | | |
|---|---|---|---|---|
| MR method | nsnp | b | se | pval |
| MR Egger | 35 | -0.15 | 0.07 | 0.047 |
| Weighted median | 35 | -0.10 | 0.05 | 0.029 |
| Inverse variance weighted | 35 | -0.14 | 0.03 | 7.76E-06 |
| Simple mode | 35 | -0.05 | 0.10 | 0.612 |
| Weighted mode | 35 | -0.08 | 0.06 | 0.218 |
| | Estimate | se | pval | |
| MR Egger - Intercept | 0.001 | 0.007 | 0.896 | |
| | Q | Q_df | Q_pval | |
| Maximum likelihood - Heterogeneity test | 29.4 | 34 | 0.694 | |
| MR Egger - Heterogeneity test | 29.5 | 33 | 0.640 | |
| Inverse variance weighted - Heterogeneity test | 29.6 | 34 | 0.685 | |
| | Estimate | SE | P | |
| MR-RAPS - Causal effect | -0.15 | 0.033 | 9.09E-06 | |
| MR-RAPS - Pleiotropy variance | 0 | 0 | NaN | |
| | RSSobs | Pvalue | | |
| MR-PRESSO Global Test | 31.5 | 0.680 | | |

| CAD (CARDIoGRAMplusC4D) → AVOI (MVP) | | | | |
|---|---|---|---|---|
| MR method | nsnp | b | se | pval |
| MR Egger | 32 | -0.20 | 0.11 | 0.091 |
| Weighted median | 32 | -0.11 | 0.07 | 0.149 |
| Inverse variance weighted | 32 | -0.14 | 0.05 | 5.69E-03 |
| Simple mode | 32 | -0.12 | 0.13 | 0.368 |
| Weighted mode | 32 | -0.16 | 0.08 | 0.053 |
| | Estimate | se | pval | |
| MR Egger - Intercept | 0.006 | 0.011 | 0.566 | |
| | Q | Q_df | Q_pval | |
| Maximum likelihood - Heterogeneity test | 32.8 | 31 | 0.378 | |
| MR Egger - Heterogeneity test | 32.6 | 30 | 0.342 | |
| Inverse variance weighted - Heterogeneity test | 32.9 | 31 | 0.372 | |
| | Estimate | SE | P | |
| MR-RAPS - Causal effect | -0.136 | 0.056 | 0.015 | |
| MR-RAPS - Pleiotropy variance | 1.66E-04 | 2.23E-04 | 0.455 | |
| | RSSobs | Pvalue | | |
| MR-PRESSO Global Test | 35.6 | 0.359 | | |

**Supplemental Table 13: MR analysis testing the effect of CAD on PCL-6 and PTSD quantitative**

| CAD (CARDIoGRAMplusC4D) → PCL-6 (UKB) | | | | |
|---|---|---|---|---|
| **MR method** | **nsnp** | **b** | **se** | **pval** |
| **MR Egger** | 33 | -0.04 | 0.08 | 0.573 |
| **Weighted median** | 33 | -0.02 | 0.04 | 0.720 |
| **Inverse variance weighted** | 33 | 0.01 | 0.03 | 0.827 |
| **Simple mode** | 33 | -0.03 | 0.08 | 0.679 |
| **Weighted mode** | 33 | -0.02 | 0.04 | 0.713 |
| | **Estimate** | **se** | **pval** | |
| **MR Egger - Intercept** | 0.006 | 0.008 | 0.463 | |
| | **Q** | **Q_df** | **Q_pval** | |
| **Maximum likelihood - Heterogeneity test** | 47.7 | 32 | 0.037 | |
| **MR Egger - Heterogeneity test** | 46.9 | 31 | 0.034 | |
| **Inverse variance weighted - Heterogeneity test** | 47.7 | 32 | 0.037 | |
| | **Estimate** | **SE** | **P** | |
| **MR-RAPS - Causal effect** | -0.000483 | 0.034 | 0.989 | |
| **MR-RAPS - Pleiotropy variance** | 1.22E-04 | 8.78E-05 | 0.166 | |
| | **RSSobs** | **Pvalue** | | |
| **MR-PRESSO Global Test** | 60.7 | 0.013 | | |

| CAD (MVP) → PTSD quantitative (PGC) | | | | |
|---|---|---|---|---|
| **MR method** | **nsnp** | **b** | **se** | **pval** |
| **MR Egger** | 46 | 0.00 | 0.03 | 0.913 |
| **Weighted median** | 46 | 0.01 | 0.01 | 0.616 |
| **Inverse variance weighted** | 46 | 0.00 | 0.01 | 0.697 |
| **Simple mode** | 46 | 0.01 | 0.03 | 0.855 |
| **Weighted mode** | 46 | 0.00 | 0.02 | 0.837 |
| | **Estimate** | **se** | **pval** | |
| **MR Egger - Intercept** | 0.001 | 0.002 | 0.754 | |
| | **Q** | **Q_df** | **Q_pval** | |
| **Maximum likelihood - Heterogeneity test** | 74.1 | 45 | 0.004 | |
| **MR Egger - Heterogeneity test** | 73.9 | 44 | 0.003 | |
| **Inverse variance weighted - Heterogeneity test** | 74.1 | 45 | 0.004 | |
| | **Estimate** | **SE** | **P** | |
| **MR-RAPS - Causal effect** | 0.010 | 0.011 | 0.378 | |
| **MR-RAPS - Pleiotropy variance** | 5.14E-06 | 4.64E-06 | 0.268 | |
| | **RSSobs** | **Pvalue** | | |
| **MR-PRESSO Global Test** | 82.2 | 0.006 | | |

**Supplemental Table 5: PRS analysis (threshold P<5E0-8) testing the association of cardiovascular outcomes and risk factors with PCL17.**

| Training | Target | r2 | P |
|---|---|---|---|
| Heart Failure | PCL-17 (MVP) | 9.40E-06 | 0.001 |
| High Blood Pressure | PCL-17 (MVP) | 1.05E-08 | 0.460 |
| Resting Heart Rate | PCL-17 (MVP) | 2.41E-06 | 0.146 |

**Supplemental Table 16: MR analysis testing the effect of heart failure on PCL-17 (MVP).**

| MR method | nsnp | b | se | pval |
|---|---|---|---|---|
| MR Egger | 8 | -2.00 | 3.76 | 0.615 |
| Weighted median | 8 | -1.35 | 0.40 | 0.001 |
| Inverse variance weighted | 8 | -1.35 | 0.36 | 1.72E-04 |
| Simple mode | 8 | -0.75 | 1.83 | 0.693 |
| Weighted mode | 8 | -1.37 | 0.37 | 0.007 |

| | Estimate | se | pval |
|---|---|---|---|
| MR Egger - Intercept | 0.071 | 0.412 | 0.869 |

| | Q | Q_df | Q_pval |
|---|---|---|---|
| Maximum likelihood - Heterogeneity test | 0.17 | 7 | 0.999 |
| MR Egger - Heterogeneity test | 0.15 | 6 | 0.999 |
| Inverse variance weighted - Heterogeneity test | 0.17 | 7 | 0.999 |

| | Estimate | SE | P |
|---|---|---|---|
| MR-RAPS - Causal effect | -2.06 | 0.226 | 7.59E-20 |
| MR-RAPS - Pleiotropy variance | 0 | 0 | NaN |

| | RSSobs | Pvalue |
|---|---|---|
| MR-PRESSO Global Test | 37.7 | 0.448 |

**Supplemental Table 17: Latent causal variable analysis testing genetic causal proportion (gcp) between PCL-17 and CAD.**

| Direction | gcp | p |
|---|---|---|
| PCL17 → CAD (UKB-CARDIoGRAMplusC4D) | 0.46±0.36 | 0.201 |
| PCL17 → CAD (CARDIoGRAMplusC4D) | 0.44±0.36 | 0.222 |
| PCL17 → CAD (MVP) | 0.34±0.43 | 0.429 |

**Supplemental Table 18: Genome-wide significant loci identified through the pleiotropic meta-analysis conducted with ASSET**

| rsID | ASSET | | Allele1 | Allele2 | CAD (UKB-CARDIoGRAMplusC4D) | | PCL-17 (MVP) | |
|---|---|---|---|---|---|---|---|---|
| | Direction | P | | | BETA | P | BETA | P |
| rs8126001 | discordant | 3.06E-08 | C | T | 0.029 | 1.69E-03 | -0.232 | 8.54E-07 |
| rs4757144 | discordant | 2.63E-08 | A | G | 0.028 | 8.36E-04 | -0.232 | 1.48E-06 |
| rs2071382 | discordant | 1.21E-16 | T | C | 0.062 | 7.14E-13 | -0.231 | 4.29E-06 |
| rs72932588 | discordant | 1.36E-19 | G | C | 0.118 | 3.02E-19 | -0.182 | 9.87E-03 |
| rs6511720 | discordant | 7.34E-22 | G | T | 0.128 | 7.88E-22 | -0.170 | 1.84E-02 |
| rs503105 | discordant | 5.00E-09 | T | C | 0.058 | 3.54E-08 | -0.164 | 6.27E-03 |
| rs7678555 | discordant | 1.93E-08 | C | A | 0.048 | 1.43E-07 | -0.141 | 6.37E-03 |
| rs2891168 | discordant | 5.75E-102 | G | A | 0.173 | 1.28E-101 | -0.141 | 2.63E-03 |
| rs12202017 | discordant | 6.52E-14 | A | G | 0.066 | 6.02E-14 | -0.118 | 3.29E-02 |
| rs10774625 | discordant | 6.35E-14 | A | G | 0.064 | 9.22E-14 | -0.108 | 2.08E-02 |
| rs9515203 | concordant | 1.52E-09 | T | C | 0.062 | 6.48E-10 | 0.128 | 1.65E-02 |
| rs12475388 | concordant | 1.01E-08 | G | A | 0.033 | 8.39E-05 | 0.215 | 7.80E-06 |
| rs11191416 | concordant | 1.57E-09 | T | G | 0.073 | 5.58E-09 | 0.219 | 5.48E-03 |
| rs288194 | concordant | 1.64E-08 | C | T | 0.027 | 9.78E-04 | 0.239 | 4.11E-07 |
| rs34811474 | concordant | 1.35E-08 | G | A | 0.034 | 8.39E-04 | 0.281 | 7.07E-07 |
| rs34484573 | concordant | 7.08E-12 | G | A | 0.078 | 1.60E-08 | 0.304 | 2.69E-05 |
| rs58673065 | concordant | 7.84E-10 | A | G | 0.031 | 1.06E-03 | 0.318 | 5.84E-09 |
